# Temporal stability of the ventral attention network and general cognition along the Alzheimer’s disease spectrum

**DOI:** 10.1101/2020.09.02.20186999

**Authors:** Evgeny J. Chumin, Shannon L. Risacher, John D. West, Liana G. Apostolova, Martin R. Farlow, Brenna C. McDonald, Yu-Chien Wu, Andrew J. Saykin, Olaf Sporns

**Affiliations:** Department of Psychological and Brain Sciences, Indiana University, Bloomington, IN; Indiana University Network Science Institute, Bloomington, IN; Department of Radiology and Imaging Sciences, Indiana University School of Medicine (IUSM), Indianapolis, IN; Indiana Alzheimer’s Disease Research Center, IUSM, Indianapolis, IN; Department of Neurology, IUSM, Indianapolis, IN

**Author notes:** Corresponding Author Evgeny J. Chumin PhD, Psychology Building 308, 1101 E 10^th^ St, Bloomington, IN 47405.

**Keywords:** Alzheimer’s disease, functional connectivity, dynamic connectivity, modularity, networks

## Abstract

Understanding the interrelationships of clinical manifestations of Alzheimer’s disease (AD) and functional connectivity (FC) as the disease progresses is necessary for use of FC as a potential neuroimaging biomarker. Degradation of resting-state networks in AD has been observed when FC is estimated over the entire scan, however, the temporal dynamics of these networks are less studied. We implemented a novel approach to investigate the modular structure of static (sFC) and time-varying (tvFC) connectivity along the AD spectrum in a two-sample Discovery/Validation design (n=80 and 81, respectively). Cortical FC networks were estimated across 4 diagnostic groups (cognitively normal, subjective cognitive decline, mild cognitive impairment, and AD) for whole scan (sFC) and with sliding window correlation (tvFC). Modularity quality (across a range of spatial scales) did not differ in either sFC or tvFC. For tvFC, group differences in temporal stability within and between multiple resting state networks were observed; however, these differences were not consistent between samples. Correlation analyses identified a relationship between global cognition and temporal stability of the ventral attention network, which was reproduced in both samples. While the ventral attention system has been predominantly studied in task-evoked designs, the relationship between its intrinsic dynamics at-rest and general cognition along the AD spectrum highlights its relevance regarding clinical manifestation of the disease.

## 1 Introduction

The pathological progression of Alzheimer’s disease (AD) is marked by abnormal aggregation of amyloid and tau proteins, neurodegeneration as measured by volumetric reduction in gray matter on magnetic resonance imaging (MRI), cognitive dysfunction, and alterations in brain networks in resting-state and task-based functional MRI (fMRI) (Dennis and Thompson, 2014). Neuroimaging methods such as high-resolution structural MRI have been shown to be valuable disease predictors prior to onset of any cognitive deficits (Jack and Holtzman, 2013, Jack et al., 2013). While structural MRI has been extensively utilized for AD diagnosis in the clinic, fMRI has yet to achieve such utility. Functional MRI studies have aimed to stratify disease groups (de Vos et al., 2018) and to identify relationships between brain function and measures of cognition, blood-based and cerebrospinal fluid biomarkers (Veitch et al., 2019), and genetic risk factors (Quevenco et al., 2017). Recently, numerous studies have focused on linking changes in behavior and/or clinical status to alterations in patterns of structural and functional brain connectivity (Douw et al., 2019, Stam, 2014), including in AD (Tijms et al., 2013).

Connectivity data obtained from fMRI has been commonly analyzed with seed-based, independent component analysis (ICA) (Córdova-Palomera et al., 2017, Fu et al., 2019), and/or graph theory methods (Contreras et al., 2019, Dennis and Thompson, 2014). Graph theory involves the construction of a matrix representation of connectivity across the whole brain, usually defining contiguous nonoverlapping regions/nodes and quantifying their pairwise relationships as a metric of statistical dependence/association (typically Pearson correlation). This functional connectivity (FC) matrix can be subdivided into coherent functional systems or resting-state networks (RSNs) (Yeo et al., 2011, Power et al., 2011). Graph measures indexing functional integration and segregation can be computed from groupwise or individual FC matrices and statistically compared across clinical groups or related to behavioral/cognitive outcomes of interest. Studies that employ graph theory on FC in AD have shown an overall decline in the internal coherence of RSNs (particularly the default mode network (Dai et al., 2019)), a reduction in separation between RSNs (Contreras et al., 2019), and alterations in various global network properties, such as modularity (Dai et al., 2019, Pereira et al., 2016).

Modularity, also referred to as community structure, is a data-driven approach for clustering nodes of a network into groups of densely interconnected regions, or communities (Sporns and Betzel, 2016). It is an important property of brain networks, with FC modules (and their reconfiguration over time) playing an important role in normal cognitive function (Bassett et al., 2011, Jang et al., 2017, Alavash et al., 2019), and various diseases (Braun et al., 2016, Contreras et al., 2019). While this is a powerful tool for studying networks, detecting network communities poses a challenging optimization problem that requires multiple iterations to achieve comprehensive sampling of partitions across multiple spatial scales (Fortunato and Barthélemy, 2007). Alterations in community structure have been reported in AD, showing higher modularity Q values in apolipoprotein E (*APOE*) ε4 carriers (Wang et al., 2015) (single community scale and one iteration), higher FC-structural connectivity coupling in the default mode network in AD (Dai et al., 2019) (single community scale and a single community partition from 1000 interations), and loss of segregation between default mode and frontoparietal RSNs (Contreras et al., 2019) (multi-scale community structure with 10,000 partitions summarized into a co-assignment matrix). Despite methodological differences in the application of modularity among these reports, they suggest a common pattern of specific disturbances of FC community structure in AD during resting-state fMRI.

In recent years it has been noted that, within a scan session, significant temporal fluctuations in functional connectivity can occur (Lurie et al., 2020, Hutchison et al., 2013), and that these fluctuations contain information that is lost when FC is estimated over the entire scan duration (static FC; sFC). Time-varying functional connectivity (tvFC) aims to capture those temporal fluctuations through estimation of connectivity across discrete or partially overlapping windows (Abrol et al., 2017, Shakil et al., 2016, Allen et al., 2014). These transient fluctuations have been studied to identify functional states that differed between diagnostic groups in AD (Gu et al., 2020), although (Schumacher et al., 2021). To date, tvFC applications in AD have relied predominantly on ICA-derived FC matrices, which have shown a reduction in internetwork connectivity in AD (Schumacher et al., 2019) and healthy aging (Tian et al., 2018). Additionally, tvFC of the interaction between dorsal attention and default mode networks has been related to cognitive reserve/function in individuals with amnestic mild cognitive impairment (MCI) (Franzmeier et al., 2017), and application of tvFC in machine learning has shown promise in disease classification (de Vos et al., 2018). However, no investigations have examined the tvFC stability of RSN networks in AD. Robustness of outcomes is difficult to assess across published studies due to methodological differences that hinder generalizability. Here, we investigate the community structure of tvFC of RSN networks in a novel approach that aims to identify group differences and relationships while reducing the number of free parameters and employing a 2-sample design with the aim of identifying robust AD related alterations. Focusing on the propensity of regions to group into communities over time (which we refer to as temporal stability), we hypothesized that any meaningful alterations in tvFC dynamics would be present in both samples. Finally, we present supplementary analyses (described in section 2.7) that aid in understanding of the observed findings.

## 2 Materials and methods

### 2.1 Sample characteristics

Data were collected as part of the Indiana Memory and Aging Study (IMAS) and from participants enrolled in the Indiana Alzheimer’s Disease Research Center (IADRC) at Indiana University School of Medicine. Informed consent was obtained from all participants or their representatives, and all procedures were approved by the Indiana University Institutional Review Board in accordance with the Belmont Report. Valid datasets from 161 participants were included in the study, consisting of 55 cognitively normal (CN; 68.24 ± 8.97 years old), 47 with subjective cognitive decline (SCD; 69.23 ± 10.80 years old), 35 with MCI (72.54 ± 7.31 years old), and 24 with AD (66.42 ± 11.26 years old). A subset of these data was included as part of a previous publication (Contreras et al., 2019). Prior to any analysis of the data, the dataset was randomly split into equally sized Discovery (n=80) and Validation (n=81) samples. Additionally, for group comparisons, the authors responsible for analysis were blinded to diagnostic group status of the participants and instead provided with neutrally coded group labels. Demographic characteristics and diagnostic group distributions for both samples are presented in Table 1. Demographic differences were assessed with an analysis of variance (ANOVA) or Χ^2^-test, where appropriate.

**Table 1.** Sample Demographics

|  | Sample |  |
| --- | --- | --- |
|  | Discovery | Validation |
| Age (years) | 69.4 ± 8.9 | 69.0 ± 10.5 |
| Education (years) | 16.3 ± 2.4 | 16.1 ± 2.7 |
| Sex (M/F) | 24/56 | 30/51 |
| Diagnostic Group (CN/SCD/MCI/AD) | 29/19/21/11 | 26/28/14/13 |

Data are shown as mean ± standard deviation. There were no significant differences between samples. M: Male; F: Female; CN: Cognitively Normal; SCD: Subjective Cognitive Decline; MCI: Mild Cognitive Impairment; AD: Alzheimer's disease.

### 2.2 Image acquisition

All participants were scanned on a Siemens 3T Prisma Scanner with a 64-channel head coil (Siemens, Erlangen, Germany). A high-resolution, T1-weighted, whole-brain magnetization prepared rapid gradient echo (MP-RAGE) volume was first acquired with parameters optimized for the Alzheimer’s Disease Neuroimaging Initiative (ADNI; http://adni.loni.usc.edu): 220 sagittal slices, GRAPPA acceleration factor of 2, voxel size 1.1 × 1.1 × 1.2 mm^3^, 5 min 12 sec duration. Resting-state functional MRI (rs-fMRI) data were acquired with a gradient-echo echo-planar imaging sequence with a multi-band factor of 3, scan time of 10 min 7 sec, and temporal resolution (TR) of 1.2 sec, resulting in 500 timepoints. Other relevant parameters were TE=29 ms, flip angle 65°, 2.5 × 2.5 × 2.5 mm^3^ voxel size, and 54 interleaved axial slices. During the scan, participants were instructed to remain still with eyes closed and to think of nothing in particular. Prior to rs-fMRI acquisition, two spin-echo echo-planar imaging (12 sec each, TR=1.56 sec, TE=49.8 ms, flip angle 90°) were acquired with reverse phase encoding directions, to be used for creating field maps for geometric distortion correction (see Supplementary Methods).

### 2.3 Image preprocessing

Data were processed with a pipeline developed in-house, implemented in Matlab (MathWorks, version 2019a; Natick, MA), and utilizing the Oxford Centre for Functional MRI of the Brain (FMRIB) Software Library (FSL version 6.0.1) (Jenkinson et al., 2012), Analysis of Functional NeuroImages (AFNI; afni.nimh.nih.gov), and ANTS (http://stnava.github.io/ANTs/) packages. This pipeline was developed and optimized for the Siemens scanner data acquired at Indiana University School of Medicine based on and following the evidence and recommendations in Satterthwaite et al. (2013), Parkes et al. (2018), and (Lindquist et al., 2019). A brief overview of preprocessing steps is provided below. For details refer to the Supplementary Methods.

All processing was carried out in each participant’s native space. T1 volumes were denoised (Coupé et al., 2008), bias field corrected (FSL), and skull stripped (ANTS). rs-fMRI data were first distortion corrected (FSL *topup*), motion corrected (*mcflirt*), and normalized to a 4D mean of 1000. Nuisance regressors were removed from the data with use of ICA-AROMA (Pruim et al., 2015), aCompCor (Muschelli et al., 2014), and global signal regression. Data were then demeaned, detrended, and bandpass filtered (0.009-0.08 Hz). Finally, 18 timepoints (∼22 sec) were removed from the beginning and end of the scan to remove edge artifacts introduced by bandpass filtering. Relative frame displacement output by *mcflirt* was used as an index of in-scanner motion.

### 2.4 Network analysis

Figure 1 illustrates a diagram of the workflow from regional time series to modularity outcomes.

**Figure 1.**
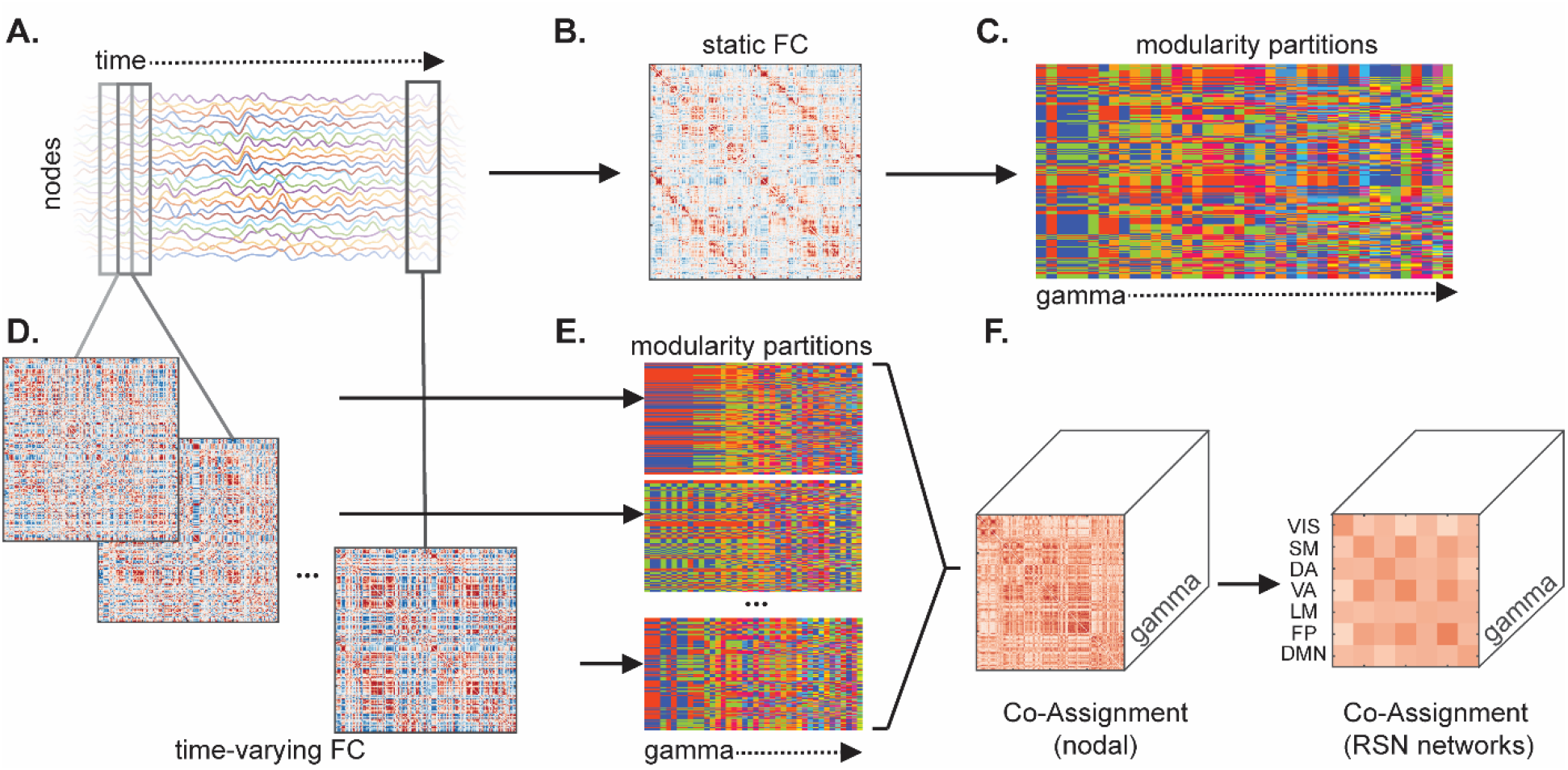
Network construction and modularity workflow. (A) Time series for all nodes were correlated to generate (B) a sFC matrix, on which modularity was performed to obtain (C) a set of partitions across the γ (gamma) parameter range. Sliding window was used to compute (D) tvFC matrices for partially overlapping time frames. (E) Modularity on each window generated a set community partitions, which were then used to compute (F) co-assignment over time at each γ. Pairwise nodal co-assignment values were then averaged with and between canonical resting state network (RSN) blocks. VIS: Visual; SM: Somatomotor; DA: Dorsal Attention; VA: Ventral Attention; LM: Limbic; FP: Frontoparietal; DMN: Default Mode Network.

#### 2.4.1 Parcellation preparation and connectivity matrix construction

For each participant, after preprocessing, the final dataset consisted of 464 timepoints. For network construction the Schaefer et al. (2018) cortical parcellations at two scales (200 and 300 regions/nodes) were used. Time series for each node were obtained from the average of all voxel time courses within that node. For sFC (the term static here refers to the assumption that FC is constant over scan duration (Lurie et al., 2020)), the full time series were cross-correlated to obtain a node-by-node matrix of Pearson correlations (Figure1 A-B). All sFC matrices were Fisher z-scored prior to analysis. The tvFC matrices were generated with a sliding window correlation approach (Figure1 A, C). This involves taking partially overlapping segments of the time series in order to estimate FC dynamics. Parameters of interest were a 56-timepoint window size (∼67 sec; above the previously reported adequate duration for tvFC estimation (Hutchison et al., 2013, Betzel et al., 2016)), step size of 14 time points (resulting in 75% overlap of adjacent windows, 30 total windows), and a taper shape of window size/3 (Zalesky et al., 2014). To investigate whether any results were dependent on window size, a longer window size (93 time point window, ∼111 sec, 28 time point steps, 14 total windows, capturing slower dynamics and mental state transitions (Iraji et al., 2020, Gonzalez-Castillo et al., 2015)) was also investigated.

#### 2.4.2 Modularity

Both sFC and tvFC matrices were analyzed with the Louvain community detection algorithm implemented in Matlab as part of the Brain Connectivity Toolbox (Rubinov and Sporns, 2010, Blondel et al., 2008), which aims to detect communities by maximizing a global modularity measure (Q-metric)

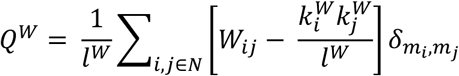

(Newman and Girvan, 2004, Rubinov and Sporns, 2010), adapted here for weighted undirected networks. To capture significant modular organization at multiple spatial scales (Jeub et al., 2018, Fortunato and Hric, 2016), we varied a partition resolution parameter γ (gamma) over a wide range, from 0.1 to 4, in steps of 0.1 (Figure 1C, E). Due to the stochastic nature of the algorithm, 1000 independent runs of modularity maximization were performed at γ value. From these 1000 runs, the single community partition with the maximal value of the Q-metric was retained. If multiple distinct partitions with equal Q-metric were identified, the one with the highest frequency across all the runs was selected. The selected partitions were then used for generating co-assignment matrices (see section 2.4.5.1 tvFC RSN temporal stability).

To maintain interpretability of the partitions obtained from modularity maximization and limit the influence of single node communities, a maximum γ value defining the upper bound of the γ range was determined for subsequent analyses. For sFC this was chosen as the highest γ at which the average number of single node communities across the sample was less than 1. The same approach was employed in tvFC, where the average was determined over all windows and all participants in the sample. Therefore, the final γ range was set within sample from 1 to the upper limit in steps of 0.1, and all analyses utilized partitions within this range.

#### 2.4.5 Network outcomes

There is mounting evidence that the community structure of brain networks is organized on multiple spatial scales (Sporns and Betzel, 2016, Betzel and Bassett, 2017). Previous clinical research which utilized Louvain modularity has either investigated a single or a small set of community scales. Here, to avoid an arbitrary selection of scale, a finely sampled range of the γ resolution parameter was assessed with a data driven selection of an upper bound for interpretable partitions. However, this yields a large set of partitions for each subject, so to identify stable variations in network outcomes, area under the curve (AUC) over the γ range was computed for outcomes of interest (sFC: Q-metric and number of communities; tvFC: mean and standard deviation of Q-metric across windows and temporal stability (defined in subsequent section)), using the trapezoidal rule as implement in the Matlab *trapz* function. If the shape of the outcome versus γ curve is not drastically different (e.g., linear versus exponential), AUC can serve as a useful composite measure, thus avoiding the need for selection of a specific scale or performing a large number of statistical tests, which introduces a multiple comparisons problem (Fornito et al., 2016).

##### 2.4.5.1 tvFC RSN temporal stability

Here temporal stability is defined as a measure that captures the variability in community structure over time (windows). Operationally, temporal stability was computed as the number of times two nodes were assigned to the same community across windows, divided by the total number of windows (co-assignment). Aggregating over all node pairs, this forms a co-assignment (CA) matrix for each γ value (Figure 1 F, left). High CA values indicate that nodes remained mostly within the same community over time, while low values indicate that nodes we often assigned to different communities. A single temporal stability matrix across γ resolutions was then computed as AUC of the CA matrices. At this stage mass univariate testing would require (N^2^/2)-N tests (where N is the number of nodes, here either 200 or 300). To reduce dimensionality of the data and improve interpretability of results, RSN-averaged temporal stability values were computed within and between bilateral RSNs (Figure 1 F, right) using the RSN labels provided with the cortical parcellations (Schaefer et al., 2018). This yields a 7×7 RSN network matrix (28 unique network blocks, 7 within network and 21 between network interaction blocks), which describes over the duration of the scan, how likely it is that nodes within a RSN or between a pair of RSNs will be assigned to the same community. It is expected that within RSN blocks will have higher temporal stability compared to between RSN blocks.

### 2.5 Neurocognitive assessments

All participants underwent a clinical and neuropsychological battery as part of the Uniform Dataset (Weintraub et al., 2018). Data from the following assessments is reported here: Montreal Cognitive Assessment (MoCA; total score) (Nasreddine et al., 2005), Rey Auditory Verbal Learning Test (RAVLT, immediate and delayed recall accuracy) (Weintraub et al., 2018), Craft Story (immediate and delayed) (Craft et al., 1996), Benson Complex Figure (presence and placement recall) (Possin et al., 2011), Trail-Making Test (time to completion) (Reed and Reed, 1997), and Digit Span Test (forward and backward number span) (Weintraub et al., 2018). All scores were adjusted by regressing out age, sex, and education (multiple linear regression in Matlab), and subsequently z-scored relative to an independent sample of controls as done in previous work (Contreras et al., 2019). Finally, the scores were grouped and averaged within three domains: (1) Global Cognition (MoCA), (2) Memory (RAVLT immediate and delayed, Craft Story immediate and delayed, Benson Complex Figure recall), (3) Attention Processing and Speed (Digit Span forward and backward, Trail A time).

### 2.6 Statistics

All statistics were carried out in Matlab. Demographic comparisons among diagnostic groups were carried out with an analysis of variance (ANOVA) or Χ^2^-test and follow-up *t*-tests, where appropriate. Group comparisons were carried out blinded to true diagnostic groups, with differences in modularity outcomes (sFC: AUCs of Q-metric and number of communities; tvFC: AUCs of means and standard deviations (across windows) of Q-metric) assessed with a permutation analysis of covariance (ANCOVA), with age, sex, and education as nuisance covariates. Temporal stability differences were probed with a permutation ANCOVA, independently for both samples and both cortical parcellation scales (28 tests per sample and parcellation). Permutations involved randomly shuffling group assignments 10,000 times to generate a null distribution of F-values, from which a permutation *p-*value was computed. Post-hoc comparisons were carried out via pairwise *t-*tests. Multiple comparisons in the were subject to a 5% False Discovery Rate (FDR) adjustment when appropriate.

Correlation analyses of modularity outcomes (Q-metric measures and number of communities) and cognitive domains were carried out within sample, unblinded, and across all groups, using Spearman’s partial correlation (adjusted for age, sex, and education). For temporal stability, correlations were investigated independently for each cognitive domain, where all 28 network blocks in the discovery sample (for both the 200 and 300 node parcellations) were tested at an uncorrected threshold of *p*<0.05. Significant relationships that emerged in both parcellations of the Discovery sample, were tested for reproducibility in the Validation sample. Multiple comparisons in the Validation sample were subject to a 5% FDR.

### 2.7 Supplementary analyses

The following analyses were carried out after the completion of the above-mentioned comparisons in order to aid interpretation of the main findings.

#### 2.7.1 Younger versus older AD

The age of AD onset has shown to have impact on neuroimaging observed deficits (Gour et al., 2014, Adriaanse et al., 2014, Chung et al., 2016). To understand if this played a role in our study, AD participants from the Discovery and Validation samples were combined and split into two subgroups based on median age at the time of the scan. The sFC and tvFC outcome comparisons were carried out at both parcellations to assess possible differences in AD subgroups due to age. Additional comparisons were carried out in CN participants to determine whether there were any differences that were age and not disease onset related.

#### 2.7.2 Impact of splitting the dataset

We investigated whether the group differences and their lack of reproducibility between samples reported here were influenced by the random split of the dataset into the Discovery and Validation samples. Modularity outcome variables for sFC (AUCs of Q-metric and number of communities) and tvFC (AUCs of the means and standard deviations of the Q-metric across windows) from both samples were pooled together and re-split 500 times, with group comparisons repeated for each split. The proportion of times (out of 500) that a result was consistent with the original split served as an indicator of robustness, with values close to 1 indicating outcomes consistent with the original split. In the case of RSN temporal stability, data were reported for each network block as the number of times (out of 500 random splits) an ANCOVA main effect of group was found in both Discovery and Validation samples, at *p*<0.05 uncorrected, within each parcellation scale.

## 3 Results

### 3.1 Sample characteristics

Table 2 shows demographic comparisons among diagnostic groups within the Discovery and Validation samples. Overall, the groups were well matched within each sample, with only the Discovery sample showing a significant difference in sex distribution (Χ^2^=8.78, *p*<0.05). Supplementary Figure 1 displays a more detailed distribution of the demographic variables, demonstrating that the sex difference in the Discovery sample is driven by greater proportions of females in the CN and AD groups. Examination of adjusted z-scores for the three cognitive domains of interest (Global Cognition, Memory, and Attention and Processing Speed) showed a significant decline with increasing diagnosis severity for Global Cognition and Memory. In-scanner motion did not significantly differ between groups (one-way ANOVA, *p>*0.05), nor was it correlated with any modularity outcome variables (static FC: AUC of Q metric or number of communities; tvFC: AUC of mean or standard deviation of Q metric or mean AUC of CA across all network blocks; *p>*0.05).

**Table 2.** Sample Characteristics

| Discovery Sample |  |  |  |  |  |
| --- | --- | --- | --- | --- | --- |
|  | CN | SCD | MCI | AD | Stats |
| N | 29 | 19 | 21 | 11 |  |
| Age (years) | 69.9±7.8 | 68.4±9.3 | 70.8±7.8 | 66.6±12.9 | n.s. |
| Education (years) | 16.5±2.4 | 16.5±2.4 | 16.3±2.5 | 15.3±2 | n.s. |
| Sex (M/F) | 4/25 | 8/11 | 10/11 | 2/9 | $p<0.05$ |
| MoCA (z-score) | -0.2±1.3‡§ | -0.4±0.9‡§ | -2.3±1*^§ | -6.3±2*^‡ | $p<0.05E-15$ |
| Memory (z-score) | -0.2±0.7‡§ | -0.2±0.5‡§ | -2.1±0.9*^ | -2.7±1.7*^ | $p<0.05E-9$ |
| Attention and Processing Speed (z-score) | -0.1±0.6 | 0.1±0.5 | -0.3±0.7 | -0.1±1.1 | n.s. |
| Mean Frame Displacement | 0.20±0.09 | 0.21±0.10 | 0.23±0.12 | 0.29±0.23 | n.s. |
| Validation Sample |  |  |  |  |  |
|  | CN | SCD | MCI | AD | Stats |
| N | 26 | 28 | 14 | 13 |  |
| Age (years) | 66.4±9.9 | 69.8±11.8 | 75.1±5.8 | 66.2±10.2 | n.s. |
| Education (years) | 16.4±2.3 | 16.5±2.7 | 15.4±2.8 | 15.5±3.2 | n.s. |
| Sex (M/F) | 5/21 | 11/17 | 8/6 | 6/7 | n.s. |
| MoCA (z-score) | 0.1±1‡§ | -0.6±1.2‡§ | -2±1.5*^§ | -5.5±2.7*^‡ | $p<0.05E-12$ |
| Memory (z-score) | -0.1±0.8‡§ | -0.3±0.8‡§ | -1.5±0.7*^§ | -3.1±0.5*^‡ | $p<0.05E-7$ |
| Attention and Processing Speed (z-score) | -0.3±0.5 | -0.0±0.6 | -0.35±0.7 | -0.2±0.5 | n.s. |
| Mean Frame Displacement | 0.20±0.08 | 0.21±0.10 | 0.23±0.12 | 0.29±0.23 | n.s. |

Data are shown as mean ± standard deviation. P-values represent significance for a one-way analysis of variance or $\chi^2$ -test where appropriate. For distributions of sex see Supplementary Figure 1. CN: Cognitively Normal; SCD: Subjective Cognitive Decline; MCI: Mild Cognitive Impairment; AD: Alzheimer's disease; n.s.: non-significant; MoCA: Montreal Cognitive Assessment; M: Male; F: Female. The following denote significant pairwise difference in post-hoc $t$ -tests: \* significantly different from CN; ^ significantly different from SCD; ‡ significantly different from MCI; § significantly different from AD.

### 3.2 Group comparisons of modularity outcomes

Modularity and correlation results (subsequent section) are reported for the 200 node parcellation and ∼1 min window. Results from the 300 node parcellation and longer window length, which were generally in agreement with the main results, are provided in supplementary figures. For sFC, the upper γ value that had on average < 1 single-node community was similar for both samples and parcellation scales (Discovery sample: γ=2.7 and 2.8 for the Schaefer 200 and 300 node networks, respectively; Validation sample: 2.8 and 2.3). Q-metric and number of communities versus γ curves were qualitatively similar (Figure 2), with their AUC values not significantly different between groups in either sample (all *p*>0.05, age, sex, and education adjusted permutation ANCOVA; 10,000 permutations). Consistent results were observed in the finer, 300 node parcellation for both samples (Supplementary Figure 2A-B).

**Figure 2.**
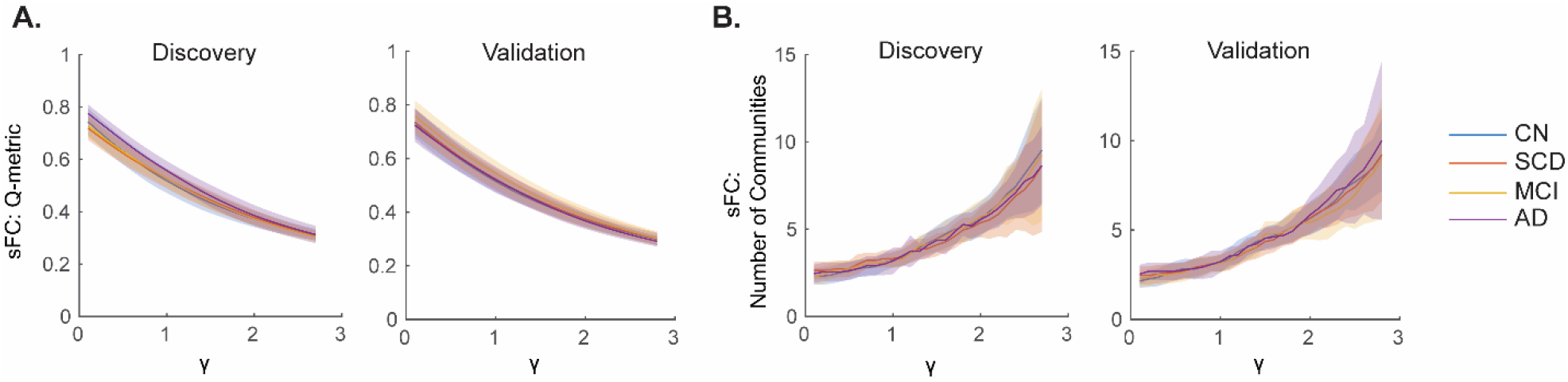
Static Functional Connectivity (sFC) modularity outcomes did not differ between diagnostic groups. Modularity (A) Q-metric and (B) number of communities versus gamma (γ) resolution curves are shown for the Discovery (Left) and Validation (Right) samples for each metric. Data are plotted as mean (solid lines) ± standard deviation (shaded fill) by group. CN: Cognitively Normal; SCD: Subjective Cognitive Decline; MCI: Mild Cognitive Impairment; AD: Alzheimer’s disease.

The average number of single-node communities across all subjects and windows for tvFC were similar to those of sFC (Discovery sample: γ=2.6 and 2.6; Validation sample: γ=2.6 and 2.2, for 200 and 300 node parcellations, respectively). Mean and standard deviation of Q-metric versus γ curves were qualitatively similar (Figure 3), with their AUC values not significantly different between groups in the Discovery sample (all *p*>0.05, age, sex, and education adjusted permutation ANCOVA; 10,000 permutations), while the Validation sample showed trend-level differences between groups for both AUC of means and standard deviations of the Q-metric (age, sex, and education adjusted ANCOVA with 10,000 permutations, *p*=0.067 and *p*=0.07). Consistent results were observed in the finer, 300 node parcellation for both samples, where the trend-level main effect of group persisted in the Validation sample (*p*=0.074 and *p*=0.069) (Supplementary Figure 2C-D). Exploratory follow-up *t*-tests revealed that the trend-level effect of group in the Validation sample was driven largely by MCI versus AD for AUC of means and SCD versus AD for AUC of standard deviations of Q-metric.

**Figure 3.**
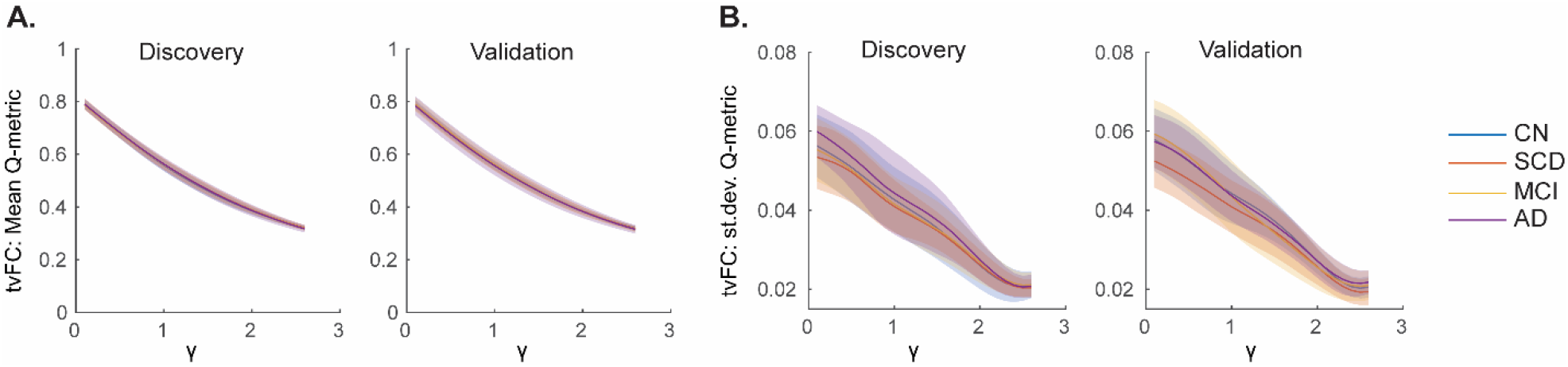
Time-varying Functional Connectivity (tvFC) modularity outcomes did not differ between diagnostic groups. Modularity Q-metric (A) means and (B) standard deviations (st.dev.) versus gamma (γ) resolution curves are shown for the Discovery (Left) and Validation (Right) samples for each metric. Data are plotted as mean (solid lines) ± standard deviation (shaded fill) by group. CN: Cognitively Normal; SCD: Subjective Cognitive Decline; MCI: Mild Cognitive Impairment; AD: Alzheimer’s disease.

As expected, CA was on average greater within RSN networks compared to between, with CA versus γ curves qualitatively similar (Supplementary Figure 3). Whole network averaged temporal stability (AUC of CA versus γ curves) moderately, but not perfectly correlated with sFC outcomes (Q metric: Spearman’s rho = 0.49 and 0.43 in the Discovery sample and 0.59 and 0.58 in the Validation sample, for the 200 and 300 node parcellations, respectively; number of communities: Spearman’s rho = -0.55 and - 0.40 in the Discovery sample, and -0.58 and -0.40 in the Validation sample, for the 200 and 300 node parcellations), which suggests that the preservation of temporal information in tvFC metrics may offer additional information over sFC modularity metrics. Nodal group averages of temporal stability between groups (Figure 4, Supplementary Figures 4 and 5) qualitatively show that high temporal stability regions in CN are reduced along AD severity in the Discovery sample (e.g., occipital cortex; Figure 4A), while low temporal stability regions show an increase (sensory/motor cortices; Figure 4A). To separate the contribution of within and between RSN network connections, network edges were stratified as either connections within a RSN or as connecting different RSNs and their respective nodal averages were computed (Figure 4B-C). Nodal averages for the Validation sample (Supplementary Figure 4) were less pronounced between groups for the full network (Supplementary Figure 4A); however, there was a qualitative increase in between network RSN temporal stability with increasing severity of diagnosis. Data from the 300 node parcellation networks (Supplementary Figure 5) were similar to that of 200 node networks.

**Figure 4.**
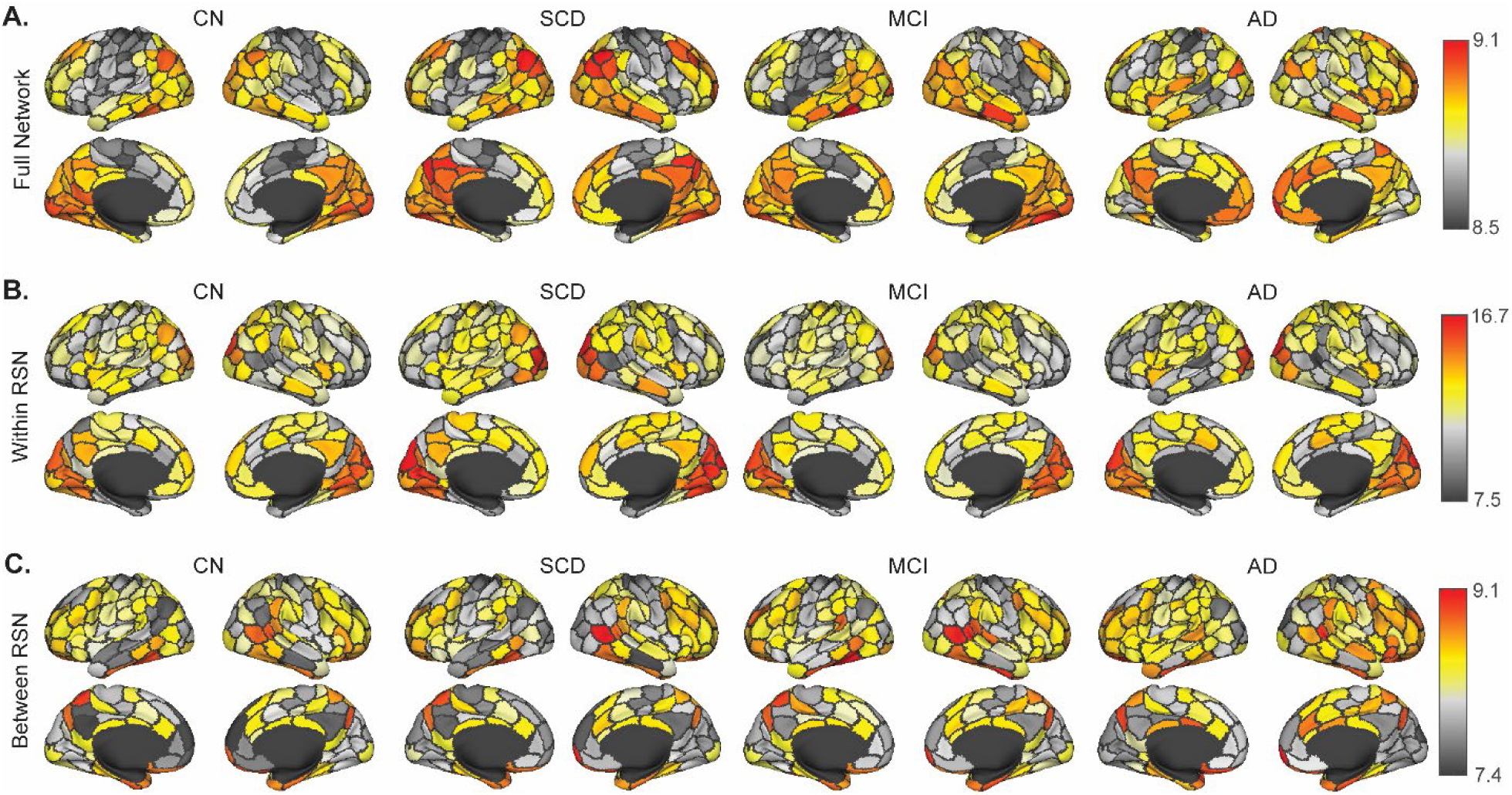
Temporal stability of each node averaging over (A) all connections of that node, (B) only the within resting state network (RSN) connections, and (C) only the between RSN connections for each node in the Discovery sample 200 node networks. Data from the Validation sample are shown in Supplementary Figure 4 for the 200 node parcellation and 300 node network data for both samples are shown in Supplementary Figure 5. CN: Cognitively Normal; SCD: Subjective Cognitive Decline; MCI: Mild Cognitive Impairment; AD: Alzheimer’s disease.

To reduce the number of multiple comparisons, statistical group differences in temporal stability were carried out on RSN block-averaged data (7 within-network and 21 between-network interactions). Several network blocks in both samples showed significant uncorrected differences (one-way permutation ANCOVA, uncorrected *p*<0.05; Figure 5B), however, there were no overlapping significant blocks between the two samples and no blocks survived FDR adjustment for the 28 tests within each sample. The main effect of group was observed in two blocks in the Discovery sample (Visual (VIS)-Somatomotor (SM) and SM-Ventral Attention (VA), Figure 5A), for which exploratory post-hoc tests revealed that AD had lower temporal stability compared to CN and SCD (*p*<0.05) for the VIS-SM interaction block, while for the SM-VA block, AD had higher temporal stability compared to the other three groups (*p*<0.05). Five blocks showed differences in the Validation sample (SM, Frontoparietal (FP), and interaction blocks of FP and SM, Limbic, and Default Mode networks, Figure 5C). Exploratory post-hoc tests showed that temporal stability was (1) higher in the SM network in MCI compared to SCD and AD, (2) lower in the SM-FP interaction block in MCI compared to SCD, (3) higher in the FP-Limbic interaction block in MCI compared to SCD and AD, (4) higher in the FP network in MCI compared to SCD and AD, and (5) higher in the FP-Default Mode interaction block in MCI compared to SCD and AD (Figure 5C; all *p*<0.05). Results from the 300 node parcellation networks were largely similar to those in Figure 5, with exception of VA-Limbic interaction block showing uncorrected significance in the Discovery sample and lack of significance in the VIS-SM interaction block (Discovery sample) and Limbic-FP interaction block (Validation sample) (Supplementary Figure 6). Finally, because there is no consensus on the window length parameter in tvFC, a longer (∼111 second) window was also employed. Both Q-metric and temporal stability results were similar for the longer window (Supplementary Figure 7).

**Figure 5.**
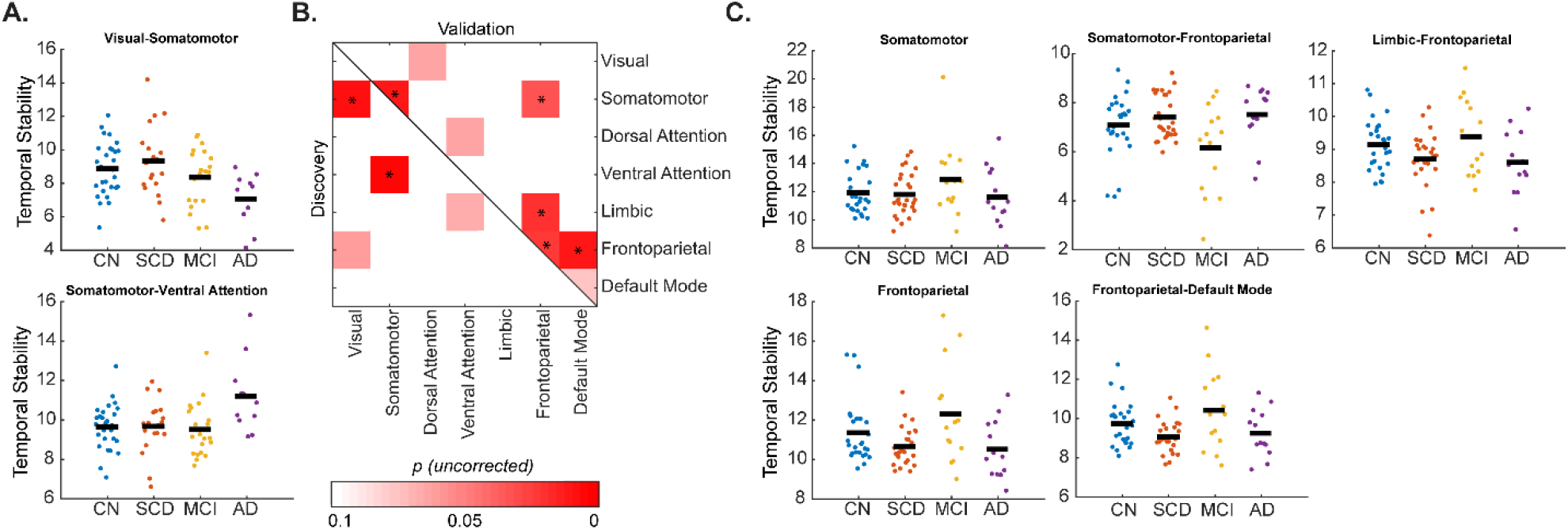
Group differences in resting state network blocks in the (A) Discovery and (C) Validation samples. (B) Permutation analysis of covariance (age, sex, and education adjusted) main effects of group within and between resting state networks at uncorrected *p*<0.05. None of the blocks were significant in both samples and none survived false discovery rate adjustment for the 28 network blocks tested. CN: Cognitively Normal; SCD: Subjective Cognitive Decline; MCI: Mild Cognitive Impairment; AD: Alzheimer’s disease.

### 3.3 Relationships to cognition

For the three cognitive domains of interest (general cognition, memory, and attention and processing speed) relationships with temporal stability were first investigated across all four diagnostic groups within all RSN blocks in the Discovery sample and any significant relationships were subsequently assessed in the Validation sample. Partial Spearman’s rho values (adjusted for age, sex, and education) are reported for all 28 RSN blocks for each of the three domains in Supplementary Figure 8. Of all investigated relationships, general cognition significantly correlated with temporal stability of the VA network (partial Spearman’s rho 0.31, *p=*0.011; Figure 6A), memory correlated with the VIS-Default mode interaction block (partial Spearman’s rho -0.27, *p=*0.032), and attention and processing speed correlated with Limbic-FP interaction block (partial Spearman’s rho 0.26, *p=*0.033). These same results were obtained in the 300 node parcellation networks (partial Spearman’s rho 0.32, -0.28, and 0.24, respectively, all *p*<0.05; Supplementary Figure 8). When these three relationships were assessed for reproducibility in the Validation sample, only the positive association of general cognition with temporal stability of the VA network was reproduced in the 200 node (partial Spearman’s rho 0.34, *p*_*FDR*_<0.05, corrected for three tests performed in the Validation sample; Figure 6B) and 300 node (partial Spearman’s rho 0.44, *p*_*FDR*_<0.05, Supplementary Figure 9A-B) parcellation networks. This relationship was also found with a longer window size (∼111 seconds) in tvFC estimation (Supplementary Figure 9C-D: 200 node parcellation, partial Spearman’s rho 0.29 and 0.33, and Supplementary Figure 9E-F: 300 node parcellation, partial Spearman’s rho 0.31 and 0.28, for Discovery and Validation samples, respectively. All *p*<0.05).

**Figure 6.**
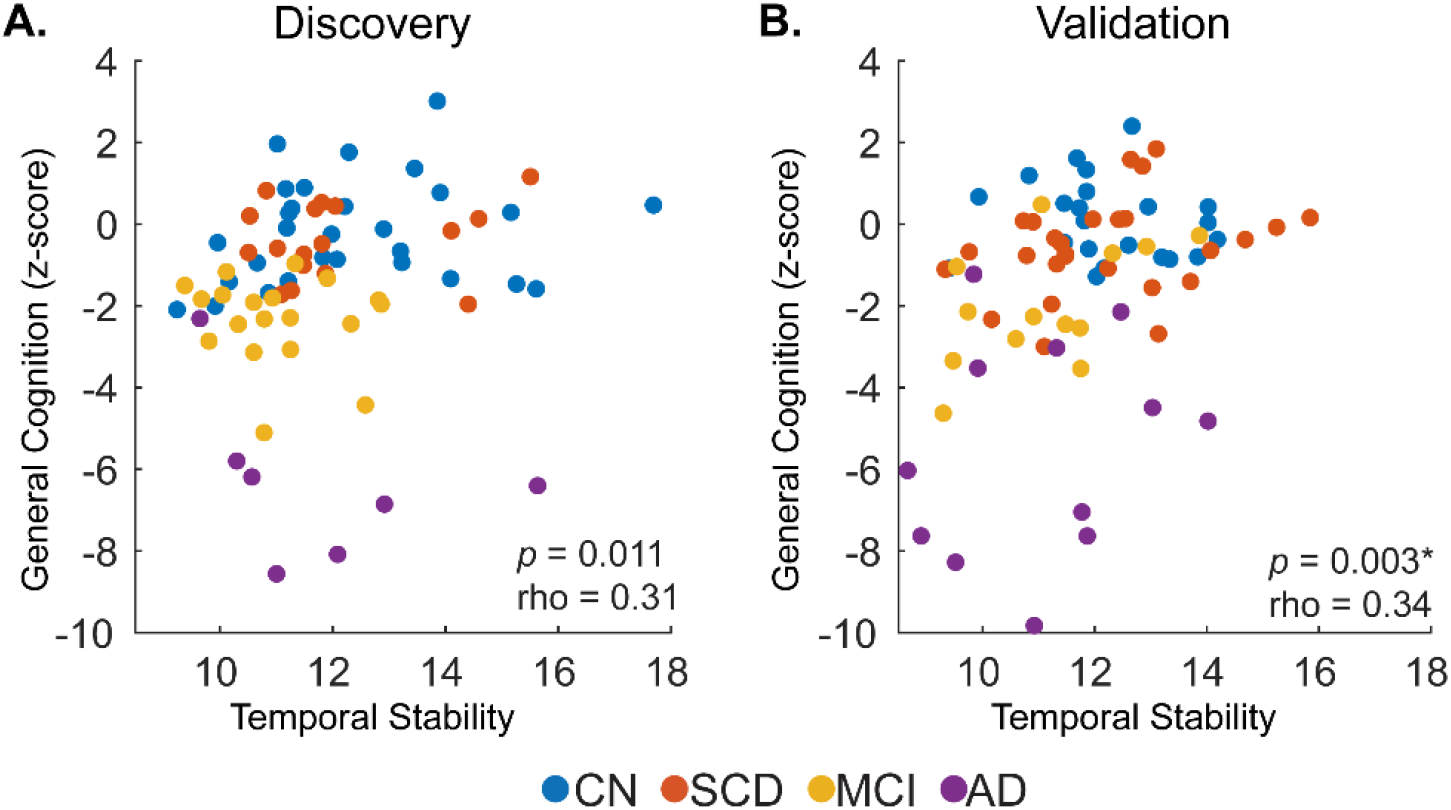
Relationship of general cognition and temporal stability of the ventral attention resting state network. Individual points represent individual participants from either the (A) Discovery or (B) Validation samples, colored by diagnostic group. CN: Cognitively Normal; SCD: Subjective Cognitive Decline; MCI: Mild Cognitive Impairment; AD: Alzheimer’s disease. rho: Partial Spearman’s rho (age, sex, and education adjusted). * denotes FDR-significant correlation in the Validation sample.

Subsequently, cognitive domains were correlated with network outcomes within each of the diagnostic groups in the combined dataset. For sFC networks, only the AUC of number of communities significantly correlated with attention and processing speed domain in the SCD group for the 200 node parcellation (partial Spearman’s rho -0.34, *p*<0.05 uncorrected), however this relationship was not present in the 300 node parcellation. For tvFC, attention and processing speed significantly correlated with mean and standard deviation of the average Q-metric over windows in the SCD group for the 200 node parcellation (partial Spearman’s rho 0.46 and -0.33, *p*<0.05, respectively), as well as with standard deviation of the average Q-metric over windows in the MCI group (partial Spearman’s rho 0.51, *p*<0.05). None of these relationships reproduced in the 300 node parcellation network modularity outcomes. Of the correlations that met uncorrected significance in the 200 node parcellation, two survived FDR adjustment in the 300 node parcellation data. Temporal stability correlated with: (1) general cognition in the CN group in the SM-VA interaction block (partial Spearman’s rho -0.39 and -0.40, *p*=0.006 and *p*_*FDR*_<0.05, for 200 (Figure 7A) and 300 (Supplementary Figure10A) node parcellation data, respectively) and (2) attention and processing speed in the MCI group in the VIS-VA interaction block (partial Spearman’s rho 0.43 and 0.50, *p*=0.022 and *p*_*FDR*_<0.05, for 200 (Figure 7B) and 300 (Supplementary Figure 10B) node parcellation data, respectively).

**Figure 7.**
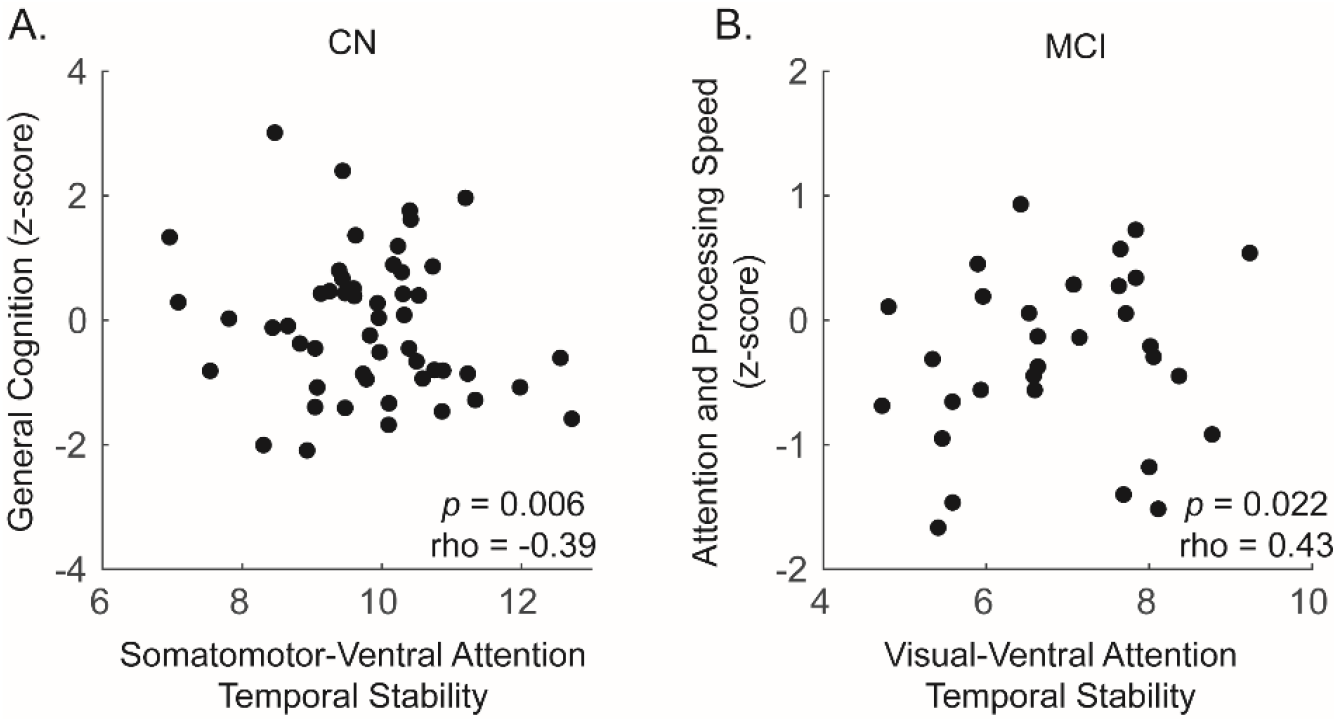
Significant within diagnostic group relationships (across both samples) between cognitive scores and temporal stability. (A) In the cognitively normal (CN) group, general cognition was negatively correlated with temporal stability of the Somatomotor and Ventral attention network interaction block. (B) In the mild cognitive impairment (MCI) group, attention and processing speed positively correlated with temporal stability of the Visual and Ventral Attention network interaction block. Data are shown for the 200 node parcellation data; 300 node parcellation data are shown in Supplementary Figure 10. Significance was determined as *p*<0.05 (uncorrected) partial Spearman’s correlation (age, sex, and education adjusted) in 200 node data that was reproduced at *p*_*FDR*_<0.05 in the 300 node data.

### 3.4 Supplementary Analyses

#### 3.4.1 Younger versus older AD comparisons

The median age of all AD participants used to separate the younger and older AD (yAD and oAD, respectively) was 65.5 years. Neither years of education nor sex differed between the two AD groups. Among modularity outcomes in sFC (AUC of modularity Q-metric and number of communities), the Q-metric was significantly lower (*t*-test, *p*<0.05) in oAD (11.74 ± 1.48) than in yAD (12.96 ± 0.99) in the 200 node but not the 300 node parcellation networks (Figure 8A). No significant differences were found for tvFC in the mean or standard deviation of the average Q-metric over windows. Temporal stability of community structure was significantly higher in the yAD group (independent samples *t*-test, *p*_*FDR*_<0.05) only in the 200 node parcellation networks for the VIS network block, the SM-Limbic interaction block, and the VA-Limbic interaction block (Figure 8 C-D). When the combined CN sample was split by age, only temporal stability was different, however, it was not in the same RSN blocks as AD (200 node parcellation, limbic network and the somatomotor-dorsal attention interaction block, independent samples *t*-test, *p*_*FDR*_<0.05).

**Figure 8.**
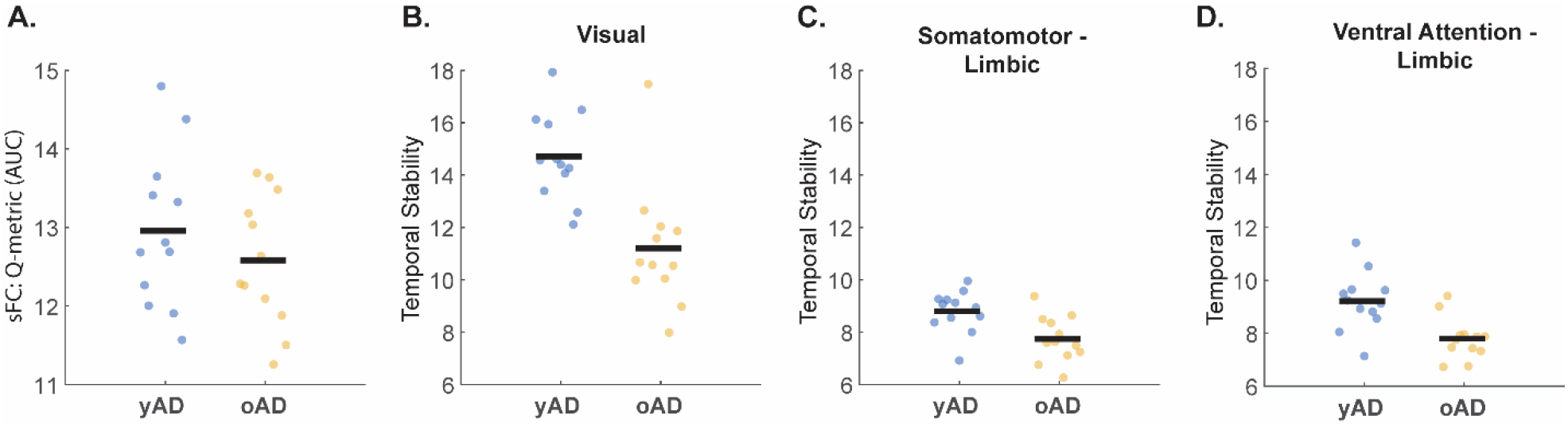
Differences in modularity outcomes in younger (yAD) compared to older Alzheimer’s disease (oAD) participants for the 200 node parcellation networks. (A) Area under the curve (AUC) of the modularity Q-metric from static functional connectivity (sFC). (B-D) Temporal stability differences of time-varying functional connectivity in the Visual network (B), Somatomotor-Limbic interaction block (C), and Ventral Attention-Limbic interaction block. A-D were significantly different between groups (independent samples *t-*tests (A) *p*<0.05 and (B-D) *p*_*FDR*_<0.05 corrected for the 28 network blocks).

#### 3.4.2 Impact of Splitting the Dataset

Repeated splits of the dataset into two samples produced results in line with those reported above. Supplementary Table 1 shows the reproducibility of findings obtained from the ‘original’ split of the dataset expressed as a ratio (# of replicated results/500 total splits). Additionally, the consensus between samples (# of times the two samples produced the same result/500) is shown. For temporal stability, in both parcellations the FP network block had the highest reproducibility of significance (0.026 and 0.076, for the 200 and 300 node parcellations, respectively), however these values were still extremely low. Across both samples and parcellations the fraction of significant outcomes of the FP network block was 0.018 (9/500 random splits; Supplementary Figure 11).

## 4 Discussion

In recent years, functional neuroimaging has been increasingly utilized to study the consequences of AD in vivo, resulting in new insights into location and extent of functional disruptions in the brain. The application of network science has allowed analysis of fMRI data as a system (network) of individual elements (brain regions) and their interactions (correlation of blood-oxygen-level-dependent signal). In this study, we focused on the temporal dynamics of brain function in AD by examining the stability of community structure, at rest, in a sample of four groups that span the AD severity continuum. In a robust, two-sample (Discovery and Validation) design, we identified a reproducible relationship of temporal stability within the ventral attention network and overall general cognition measured by the MoCA. Lower stability, measured as a weakened propensity of ventral attention network regions to associate within the same network community across time, was significantly associated with lower levels of performance. We found no persistent/reproducible differences in commonly reported modularity outcomes (Q-metric, its mean and standard deviation over time, or the number of communities).

Prior applications of modularity analyses vary with respect to the algorithm used to cluster the data. The two most often seen approaches utilize either the Louvain algorithm (Blondel et al., 2008, Rubinov and Sporns, 2010, Contreras et al., 2019, Brier et al., 2014, Onoda and Yamaguchi, 2013) or k-means clustering (Ma et al., 2020, Schumacher et al., 2019). When investigating the community structure of sFC networks (computed via average over total scan duration), we did not find any group differences when using the area under the curve (AUC) of Q-metric or number of communities over the γ resolution range. Other studies that utilized the Louvain algorithm have shown a relationship of the Q-metric with age for a predefined community partition (Brier et al., 2014), for a single γ value (Onoda and Yamaguchi, 2013), and for multiscale modularity outcomes (Contreras et al., 2019) over the full γ range. To our knowledge, no studies have utilized AUC to assess modularity differences that are consistent over a range of γ values. AUC can be a robust outcome measure for metrics that are dependent on free parameters (i.e., the γ resolution parameter in modularity), as it can capture behavior over a range of parameter values and reduce the number of statistical comparisons. The absence of group differences in AUC of the Q metric and number of partitions reported here highlights the importance of methods that sample a range of free parameters. Selection of a single or a few values can produce outcomes that are algorithm/scale dependent, while composite measures such as AUC provide a generalizable framework that can be applied across datasets.

Either during performance of a task or at rest, blood-oxygen-level-dependent signal is always fluctuating, in part due to moment-to-moment changes in neuronal activity. It is now generally accepted that these temporal dynamics are meaningful for our understanding of brain function (Hutchison et al., 2013, Allen et al., 2014). Studies have shown that brain FC fluctuates between segregated and integrated states (Fukushima et al., 2018), that tvFC is related to cognition (Cohen, 2018, Kucyi et al., 2018), and that variance in tvFC may be genetically influenced (Barber et al., 2021). Time-varying FC is often quantified via a sliding window approach, where partially overlapping segments of data are used to study temporal changes in outcomes of interest. In AD, this method has shown that patient groups spend more time in a connectivity state characterized by weak correlations (Fu et al., 2019, Schumacher et al., 2021). These studies utilized a k-means clustering procedure to identify these states and did not directly probe the modular structure of those networks. Here we used the Louvain algorithm to assess variability in modularity (AUC of the Q-metric), similar to the approach utilized in Hilger et al. (2020). We found no difference in average or standard deviation of the Q-metric across diagnostic groups. Visual inspection of the curves in Figure 3 shows the group averages overlapping one another over the full γ range, indicating that the lack of difference is not due to AUC computation.

Hilger et al. (2020) referred to temporal stability as the variance in the modularity Q-metric. Here we used Co-Assignment (CA) (Jeub et al., 2018) as an index of temporal stability, similar to Contreras et al. (2019), in which CA was computed from partitions across spatial scales to investigate coherence and coupling of RSNs in static FC. Because CA describes a relationship among node pairs, it makes it distinct from other tvFC network metrics, such as flexibility, which is a single node-based metric that relies on multi-layer modularity approaches (Bassett et al., 2011). We computed the AUC of CA over time (across all sliding windows) to investigate stability of community assignments within and between cortical RSNs. For 28 unique cortical RSN blocks (7 within network and 21 interaction blocks), commonly implicated networks emerged when group comparisons were carried out (Figure 5, Supplementary Figure 6, and Supplementary Figure 7 for longer window size), such as default mode and frontoparietal involving systems (Hohenfeld et al., 2018). However, our findings did not replicate in our two-sample design and none of the differences survived FDR-adjustment for the 28 network blocks.

We assessed whether the random split of the dataset into Discovery and Validations samples influenced our outcomes by performing an additional 500 random splits and repeating all statistical comparisons on the outcome metrics. Results from this analysis demonstrated a high degree of consensus with the original dataset split (Supplementary Table 1 and Supplementary Figure 11), which indicates that the absence of reproducibility of group differences (or lack thereof) in our study is not a chance occurrence. It has been shown that FC differs as a function of age of onset of AD (Pini et al., 2020); therefore, in order to assess whether our findings are influenced by a heterogeneous AD sample we performed a median split on the AD patient group based on age at time of scan and compared older and younger patients. The median age was 65.5 years, which is consistent with generally accepted criteria for classification of early and late onset Alzheimer’s disease. Our results showed that younger AD patients had higher Q-metric AUC (sFC) as well as higher temporal stability of visual, somatomotor-limbic, and ventral attention-limbic network interaction blocks. These observed differences did not overlap with any findings among our diagnostic groups, and while they are in themselves intriguing, they must be further addressed in larger samples before any definitive interpretations can be made.

Correlation analysis of RSN temporal stability and neuropsychological domains (global cognition, memory, and attention processing and speed) showed a single robust and reproducible relationship of global cognition and the ventral attention network. This relationship was seen across the two window sizes, at both parcellation scales, and in both Discovery and Validation samples (Figure 6). The ventral attention network is active during orientation to salient targets (Fox et al., 2006, Yeo et al., 2011). It has been shown to activate during a short-term memory task in AD in a manner that did not significantly differ from controls (Kurth et al., 2019), and an ICA-derived ventral attention network was shown to be preserved in AD (Li et al., 2012). The temporoparietal junction, a key node in the ventral attention network, has been shown to have altered connectivity to the posterior cingulate cortex in AD patients with poor orientation for time performance, relative to AD patients with good performance (Yamashita et al., 2019). Our index of global cognition was derived from the total Montreal Cognitive Assessment score, which captures several cognitive domains, such as memory, attention, executive function, language, and reasoning, which require attention and orientation to stimuli. Therefore, it is plausible that lower stability of the ventral attention system contributes toward observed deficits in global cognition.

There are important methodological considerations and limitations to be considered in the interpretation of the presented findings. First, data preprocessing strategies can impact the final FC estimates. The strategy employed here is one that is well suited for dynamic FC analyses and has been shown to produce reliable network estimates (Parkes et al., 2018). How the brain is parsed and in turn how regions are grouped can have an impact on the data. We chose the Schaefer parcellation as a functionally relevant subdivision of the cortex, the regions of which are discretely group into the RSNs reported by Yeo and colleagues. Alternative strategies could involve use other available parcellation, testing each node pair (this poses a large multiple comparison problem), or testing the whole cortex average (very limited spatial specificity). Additionally, tvFC and modularity require free parameter choices, such as choice of window size, number of modularity iterations, and resolution parameter range. We aimed to address these issues by choosing a shorter and longer window size, running a high number of iterations (1,000) of modularity, and utilizing area under the curve across a range of the resolution parameters. It is possible that fluctuations in tvFC are related to sampling variability in fMRI data; however, we do not believe this to be the case here, as a longer (∼1min and ∼2min) window was utilized with a taper function 1/3^rd^ the window size. Another limitation of these data is a relatively short acquisition duration of fMRI data (∼10 min). It is now generally accepted that longer acquisitions are better for more robust estimates of FC, however, this is difficult to achieve in clinical data, particularly in elderly and patient populations. Finally, splitting the dataset resulted in moderate sample sizes (∼80 participants per sample). While larger samples are always preferred in order to conduct sufficiently powered analyses, we believe the validation opportunity provided by the two-sample strategy is more advantageous for identification of robust findings.

In conclusion, we investigated modular structure of static and time-varying FC in a sample of four groups along the AD continuum. We report a robust relationship between global cognitive performance measured by MoCA and temporal stability of the ventral attention network. While FC-based metrics are not yet capable of serving as disease biomarkers in AD, time-varying FC investigations of resting-state fMRI data may offer unique insight into the neurobiological consequences of AD as well as inform clinical interventions as well as biomarker and treatment development, specifically when evaluating their impact on cognition through stability of attentional systems.

## Data Availability

Any pertinent code and anonymized data requests should be made to the corresponding author.

## 7 Author Contributions

**Evgeny Chumin:** Conceptualization, Methodology, Formal Analysis, Software, Writing-Original Draft, Writing-Review & Editing. **Shannon Risacher:** Methodology, Resources, Data Curation, Writing-Review & Editing, Funding Acquisition. **John West:** Resources, Data Curation. **Liana Apostolova:** Methodology, Writing-Review & Editing, Funding Acquisition. **Martin Farlow:** Writing-Review & Editing. **Brenna McDonald:** Writing-Review & Editing, Project Administration. **Yu-Chien Wu:** Writing-Review & Editing, Funding Acquisition. **Andrew Saykin:** Writing-Review & Editing, Supervision, Funding Acquisition. **Olaf Sporns:** Conceptualization, Methodology, Software, Writing-Original Draft, Writing-Review & Editing, Supervision, Funding Acquisition.

## 8 Acknowledgements

The authors thank Eileen Tallman, Aaron Vosmeier, Rachael Deardorff, Bradley Glazier, Kala Hall, Lili Kyurkchiyska, Evan Finley, Yolanda Graham-Dotson, Steve Brown, and Sujuan Gao for their contributions to this study. We also thank the participants in the Indiana Memory and Aging Study (IMAS) and their family members without whom this research would not be possible.

