## Supplemental methods/figs for "Temporal stability of the ventral attention network and general cognition along the Alzheimer’s disease spectrum"

**Supplementary Materials**

**Supplementary Table 1.** Impact of Dataset Split on Observed Outcomes

|  | Schaefer 200 Nodes | | | Schaefer 300 Nodes | | |
| --- | --- | --- | --- | --- | --- | --- |
|  | Sample 1 | Sample 2 | Consensus between samples | Sample 1 | Sample 2 | Consensus between samples |
| Static FC |  |  |  |  |  |  |
| Q-metric | 0.97 | 0.96 | 0.93 | 0.94 | 0.94 | 0.88 |
| Number of Partitions | 0.95 | 0.93 | 0.88 | 0.99 | 0.99 | 0.98 |
| Time-Varying FC |  |  |  |  |  |  |
| Mean Q-metric | 0.90 | 0.92 | 0.83 | 0.89 | 0.88 | 0.77 |
| Standard Deviation of the Q-metric | 0.88 | 0.88 | 0.76 | 0.83 | 0.83 | 0.66 |

Data are shown as a fraction of times that a dataset split (out of a total of 500 random splits) was consistent with results obtained in the original split. Consensus between samples is the fraction of times results from both samples in a single split were consistent with the original split. FC: Functional Connectivity

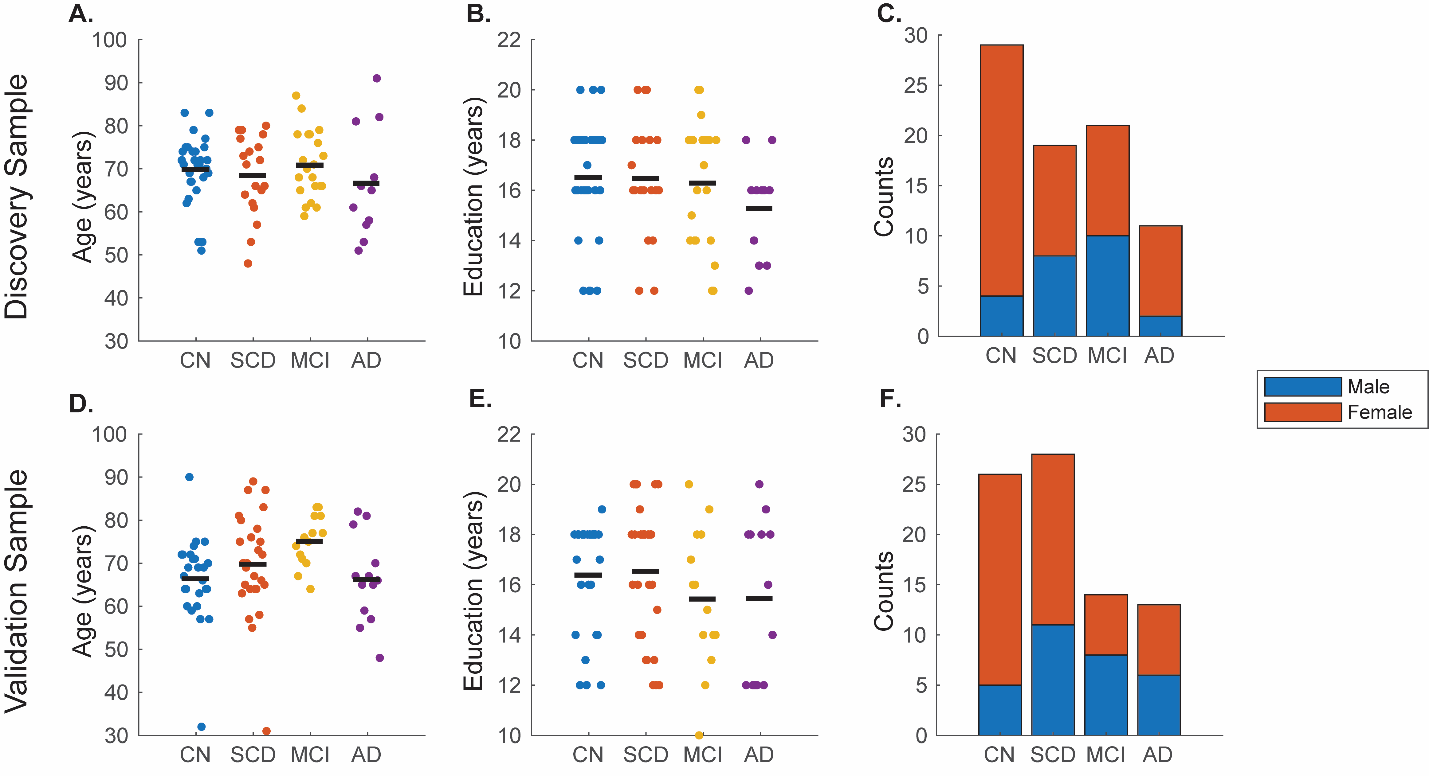

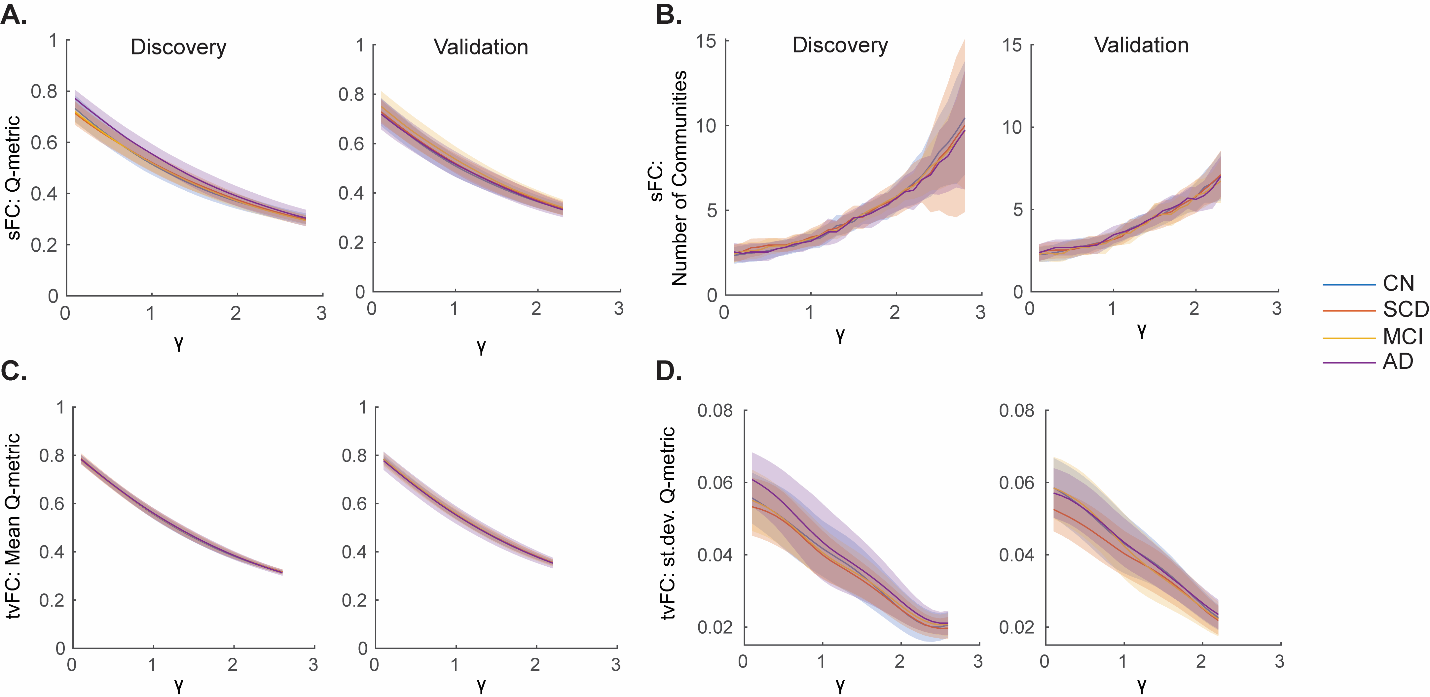

**Supplementary Figure 1.** A detailed overview of age (A, D), education (B, E), and sex (C, F) for the Discovery (Top) and Validation (Bottom) samples. Individual points in scatter plots represent individual subjects while the group means are denoted by the solid black line. Stacked boxplots (C, F) show sex distributions (female: orange and male: blue) for each sample. CN: Cognitively Normal; SCD: Subjective Cognitive Decline; MCI: Mild Cognitive Impairment; AD: Alzheimer’s disease.

**Supplementary Figure 2.** Modularity outcomes of static (sFC; A-B) and time-varying functional connectivity (tvFC; C-D) in 300 node parcellation networks. Data are plotted as mean (solid lines) ± standard deviation (shaded fill) by group. CN: Cognitively Normal; SCD: Subjective Cognitive Decline; MCI: Mild Cognitive Impairment; AD: Alzheimer’s disease; st.dev.: standard deviation.

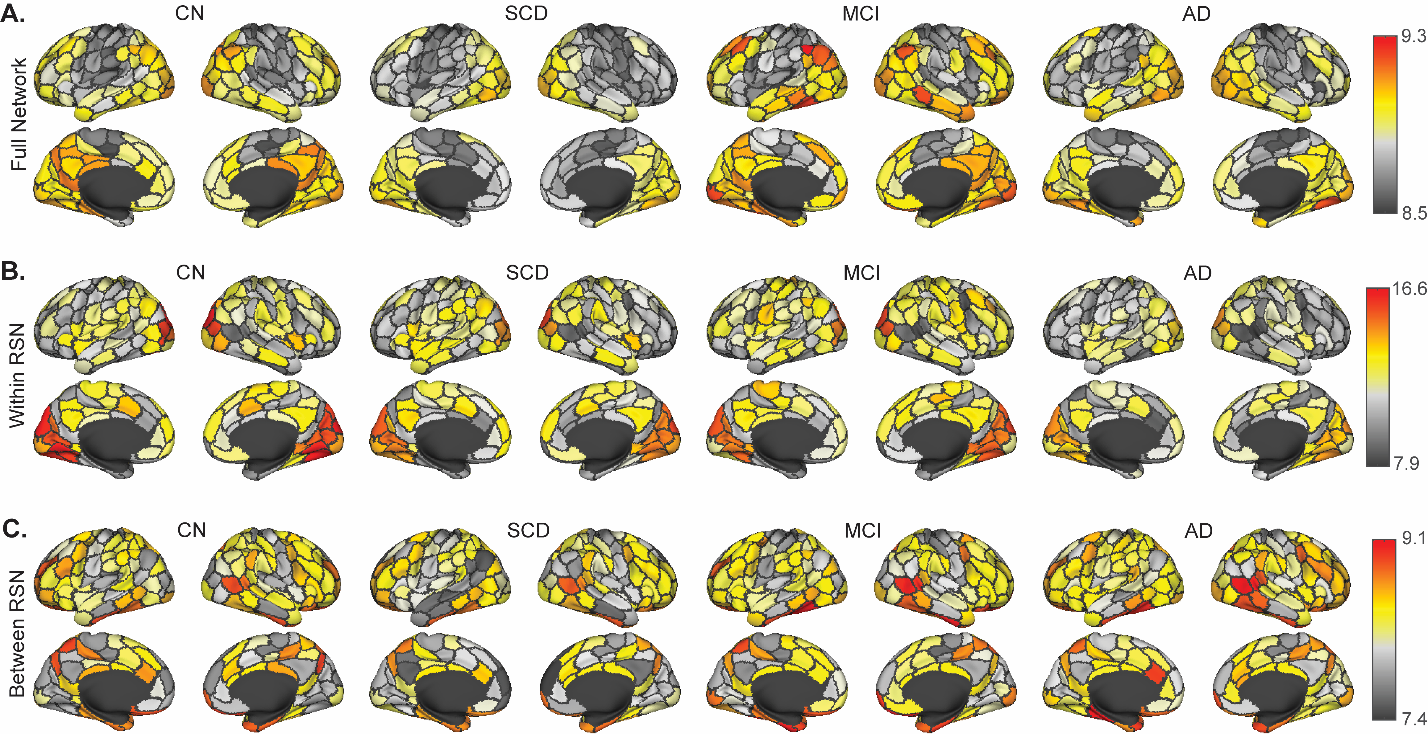

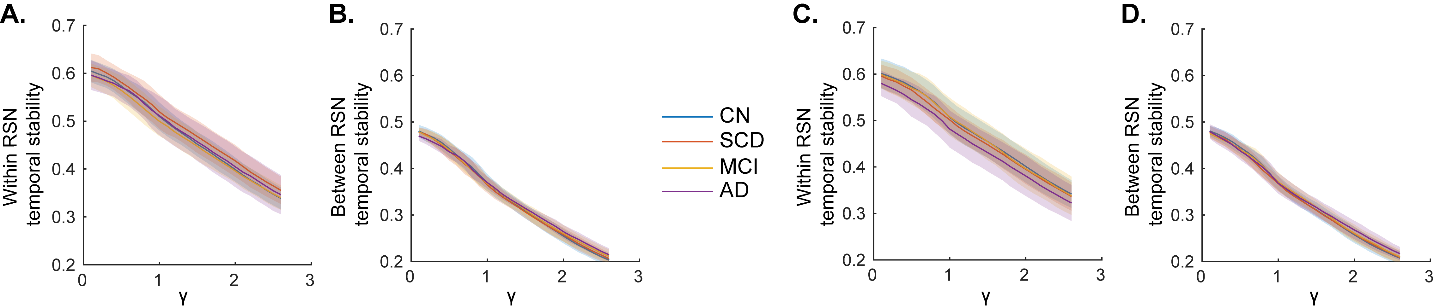

**Supplementary Figure 4.** Temporal stability of each node averaging over (A) all connections of that node, (B) only the within resting state network (RSN) connections, and (C) only the between RSN connections for each node in the Validation sample 200 node networks. CN: Cognitively Normal; SCD: Subjective Cognitive Decline; MCI: Mild Cognitive Impairment; AD: Alzheimer’s disease.

**Supplementary Figure 3.** Temporal stability averaged for within and between resting state network (RSN) blocks for (A-B) Discovery and (C-D) Validation samples. As expected within RSN values are greater than between RSN. Temporal stability versus gamma (γ) resolution curves are similar in shape, which allows for area under curve comparisons. Data are plotted for the 200 node parcellation as mean (solid lines) ± standard deviation (shaded fill) by group. CN: Cognitively Normal; SCD: Subjective Cognitive Decline; MCI: Mild Cognitive Impairment; AD: Alzheimer’s disease.

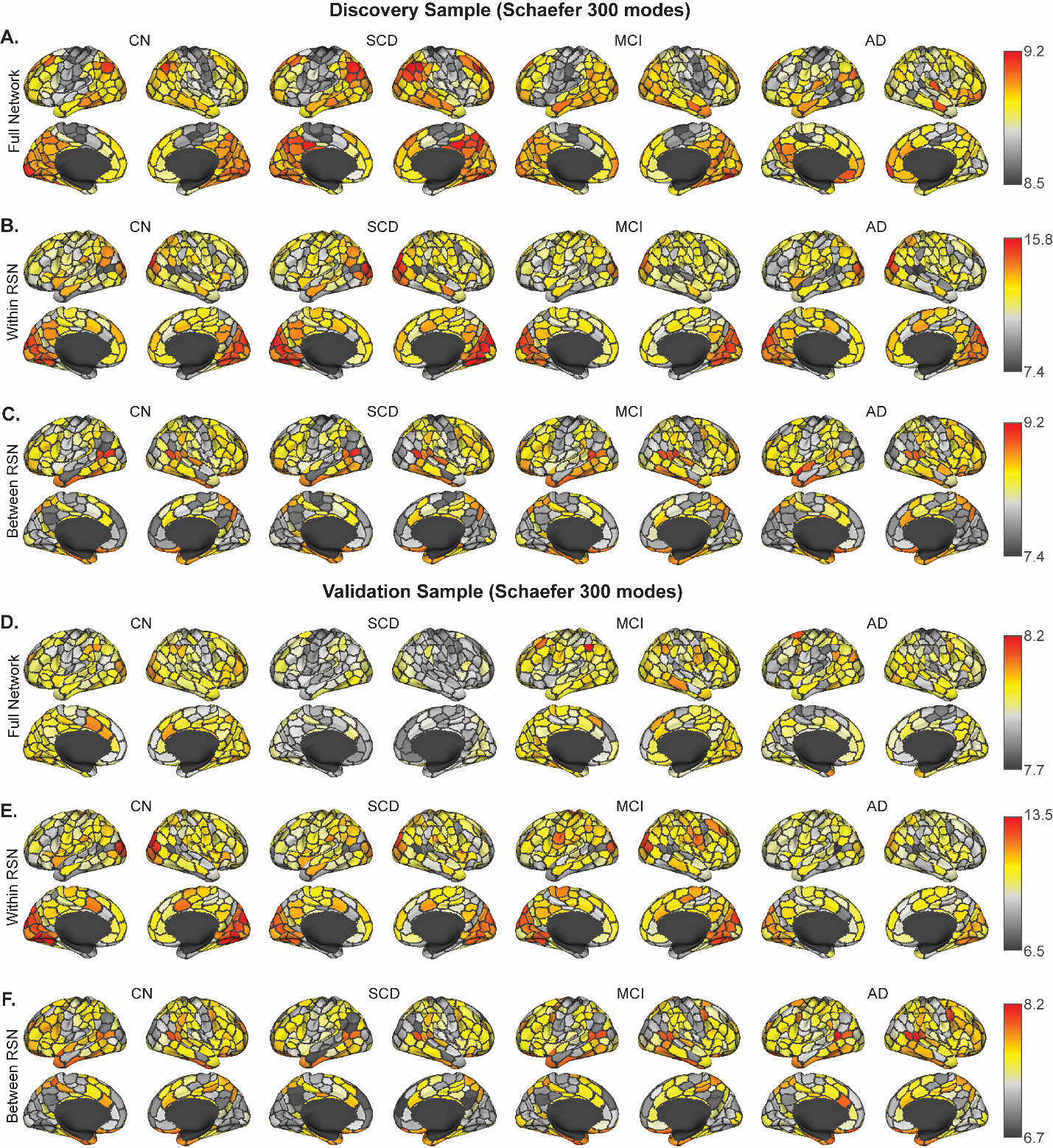

**Supplementary Figure 5.** Temporal stability of each node averaging over (A, D) all connections of that node, (B, E) only the within resting state network (RSN) connections, and (C, F) only the between RSN connections for each node in the (A-C) Discovery and (D-F) Validation sample 300 node networks. CN: Cognitively Normal; SCD: Subjective Cognitive Decline; MCI: Mild Cognitive Impairment; AD: Alzheimer’s disease.

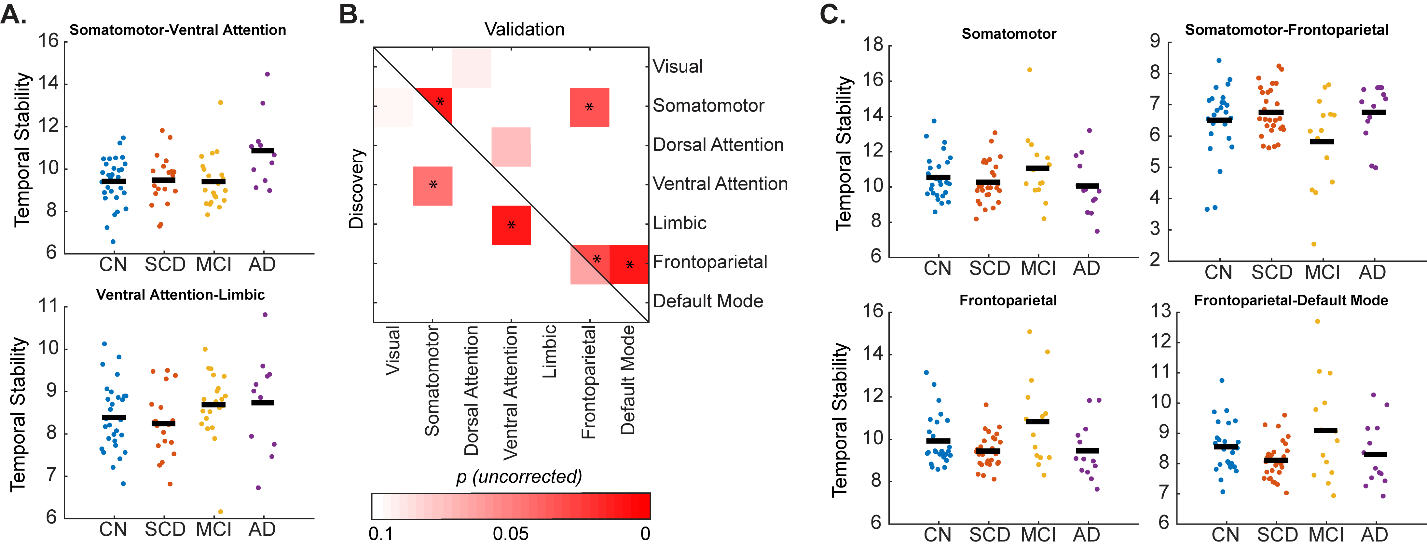

**Supplementary Figure 6.** Group differences in resting state network blocks in the (A) Discovery and (C) Validation samples in the 300 node parcellation. (B) Permutation analysis of covariance (age, sex, and education adjusted) main effects of group within and between resting state networks at uncorrected *p*<0.05 were similar to the 200 node networks. None of the blocks were significant in both samples and none survived false discovery rate adjustment for the 28 network blocks tested. CN: Cognitively Normal; SCD: Subjective Cognitive Decline; MCI: Mild Cognitive Impairment; AD: Alzheimer’s disease.

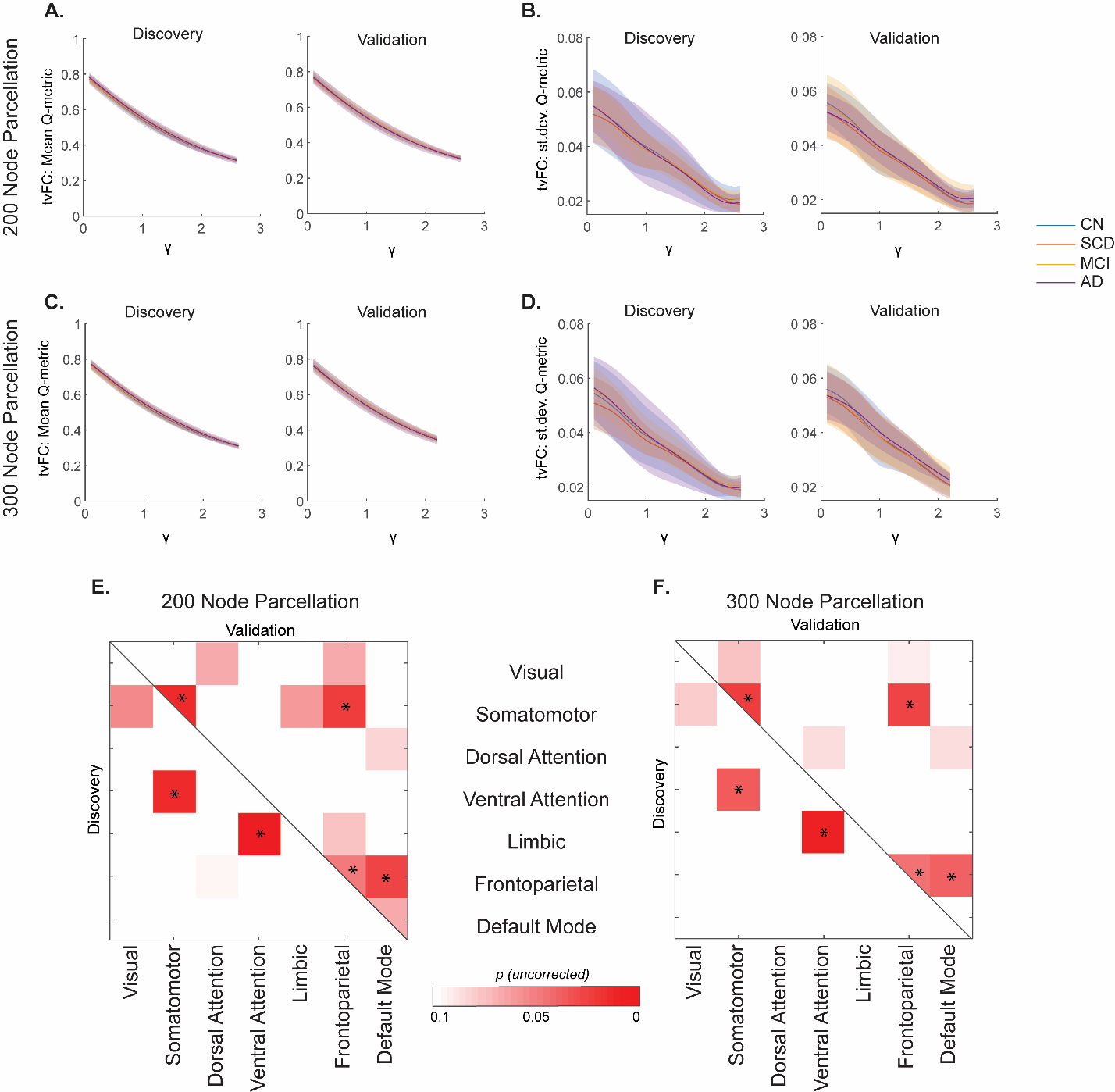

**Supplementary Figure 7.** Modularity outcomes for time-varying functional connectivity (tvFC) with a 95-time point (~111 second) window. (A-B) Mean and standard deviation of average modularity Q-metric across gamma (γ) resolution over windows in the 200 node parcellation and (C-D) 300 node parcellation. (E-F) Matrix representations of significant (*p*<0.05 uncorrected) differences temporal stability (one-way permutation ANCOVA at each network block). These results, with a longer window, are largely consistent with the ~1-minute window results reported in the main text. CN: Cognitively Normal; SCD: Subjective Cognitive Decline; MCI: Mild Cognitive Impairment; AD: Alzheimer’s disease.

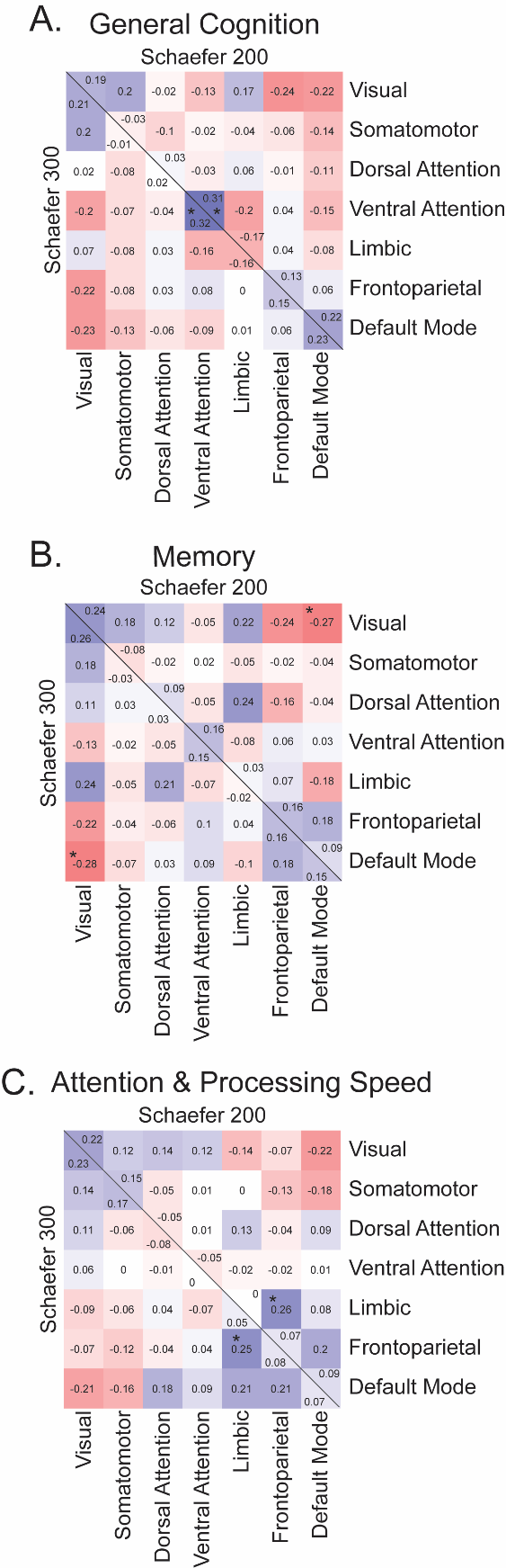

**Supplementary Figure 8.** Correlations of neuropsychological domains and resting state network block temporal stability in the Discovery sample. Colors and corresponding values denote strength/direction of the relationship (partial Spearman’s rho, adjusted for age, sex, and education). The diagonal line separated data for the 200 (upper triangular) and 300 (lower triangular) node parcellation networks. *Significant at *p*<0.05, uncorrected.

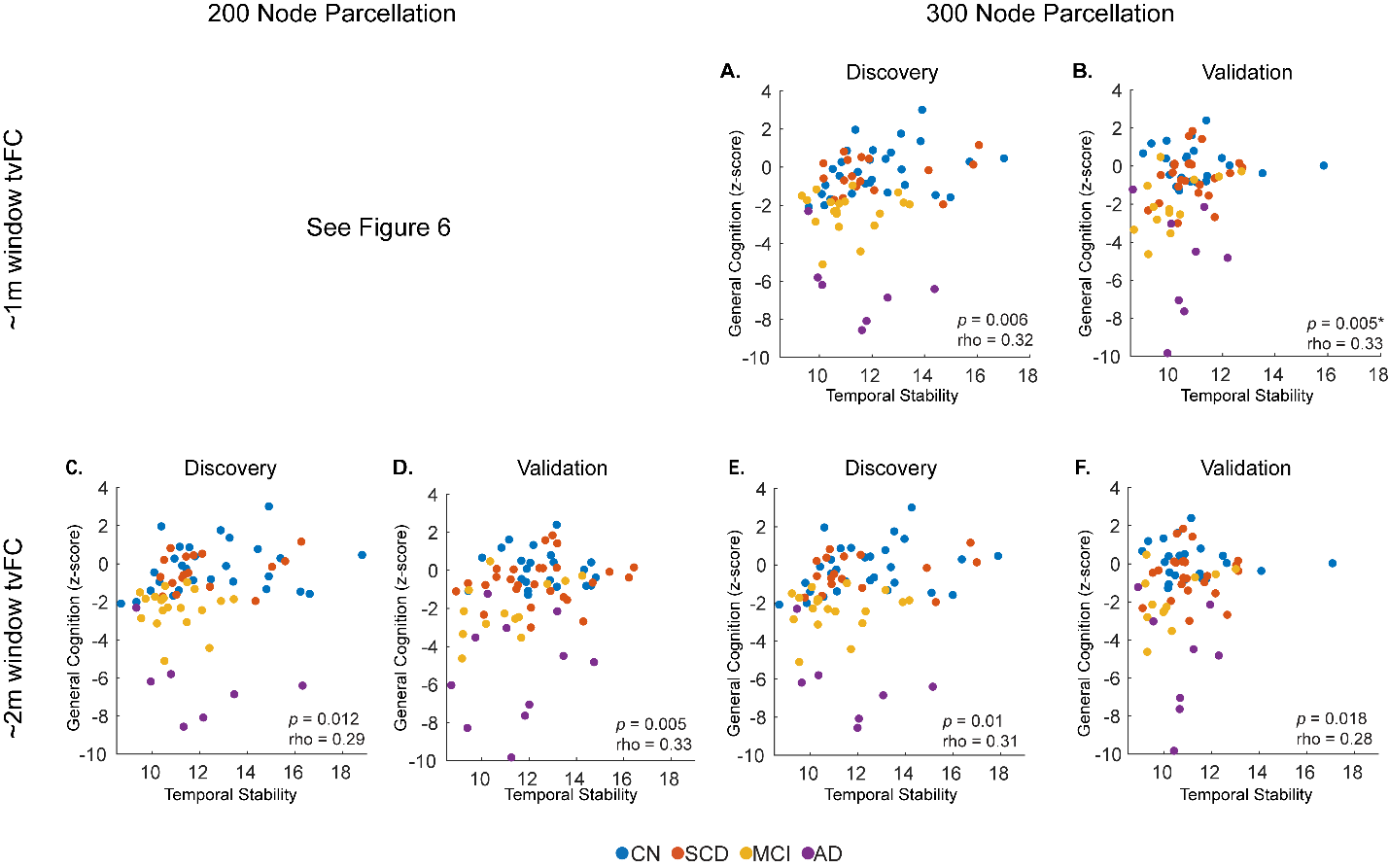

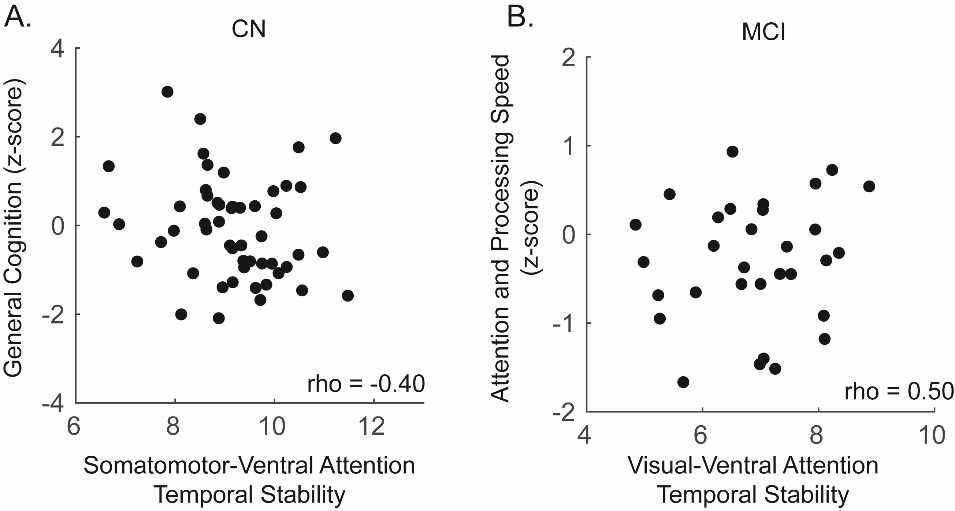

**Supplementary Figure 10.** Significant within diagnostic group relationships between cognitive scores and temporal stability in 300 node parcellation data. Data are consistent with 200 node parcellation data presented in Figure 7. Correlations are significant at *p_FDR_*<0.05. CN: Cognitively Normal, MCI: Mild Cognitive Impairment, rho: partial Spearman’s rho (age, sex, and education adjusted).

**Supplementary Figure 9.** Relationship of general cognition and temporal stability of the ventral attention resting state network in the (A-B) ~1 min window 300 node parcellation data and (C-F) ~2 min window (C-D) 200 node and (E-F) 300 node parcellation tvFC data. Individual points represent individual participants colored by diagnostic group. CN: Cognitively Normal; SCD: Subjective Cognitive Decline; MCI: Mild Cognitive Impairment; AD: Alzheimer’s Disease. rho: Partial Spearman’s rho (age, sex, and education adjusted). * denoted FDR-significant correlation in the Validation sample.

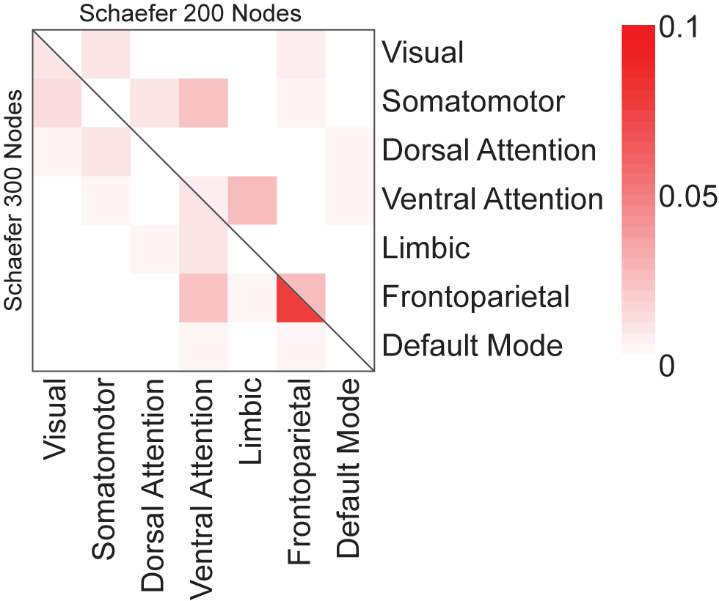

**Supplementary Figure 11.** Robustness of network block temporal stability (area under the curve of co-assignment across windows) of the original dataset split, compared to 500 additional random dataset splits, for the 200 node (lower triangle) and 300 node (upper triangle) parcellations. Rows and columns are labeled according to the canonical resting-state networks. Data are shown as the fraction of times (out of 500) a significant group difference was observed (*p*<0.05, uncorrected) for both samples in a split.

**Supplementary Methods**

**Image Processing**

**Overview.** The image processing pipeline was developed at Indiana University School of Medicine, following best practices guidelines in the neuroimaging field (Satterthwaite et al., 2013, Power et al., 2015, Parkes et al., 2018, Lindquist et al., 2019). It is a Matlab-based set of scripts, which utilize FSL (Jenkinson et al., 2012), AFNI (Cox, 1996), and ANTS (<http://stnava.github.io/ANTs/>) to take data from raw neuroimaging format to connectivity matrices, while maintaining the data in native participant space. The following sections describe each preprocessing section in detail with supporting references.

**T1 preprocessing**

*Dicom Import:* Data are first converted from raw dicom (.dcm) format to nifti (.nii) with the dcm2niiX tool (Li et al., 2016)( <https://github.com/rordenlab/dcm2niix>).

*Denoising:* For each dataset, the T1 nifti image is then denoised using an optimized nonlocal means filter for 3D MRI implemented in Matlab (Coupé et al., 2010, Coupé et al., 2008), to improve subsequent brain extraction, tissue segmentation, and registration steps.

*Bias Field Correction:* Bias field correction is done within the FSL’s FAST tool (Zhang et al., 2001) with *robustfov* field-of-view cropping.

*Brain Extraction:* Across the whole sample, first pass brain extraction was carried out with ANTS (<https://github.com/ANTsX/ANTs>) using an openly available OASIS data template (<https://figshare.com/articles/ANTs_ANTsR_Brain_Templates/915436>). All brain masks were then visually checked for quality in Matlab by visualizing 5 evenly spaced slices of the T1 with the mask as a semi-transparent overlay. For any participant with a poor quality mask, brain extraction was repeated with an alternative template from the same source that was derived from the Nathan Kline Institute (<http://dx.doi.org/10.15387/fcp_indi.corr.nki1>). For any remaining participants for which a suitable brain mask was still not attained, FSL bet with -B option was used to derive a mask, by tuning the fractional intensity and vertical gradient in fractional intensity parameters independently for each participant to obtain a suitable mask. Finally, any gaps/holes in the mask were filled with *fslmaths -fillh* option and a final brain masked image was extracted.

*Standard Space Registration:* In order to apply parcellations in native space, linear and nonlinear transformations were generated for each participant T1 to/from Montreal Neurological Institute (MNI) standard space via the following:

1. FSL FLIRT (Jenkinson and Smith, 2001) linear 6 degrees of freedom registration (dof6) to MNI.
2. FSL FLIRT affine 12 degrees of freedom registration (dof12) of dof6 to MNI.
3. FSL FNIRT (Andersson et al., 2010) nonlinear warp of dof12 to MNI

The 2 transformation matrices and warp field were then inverted with the *convert_xfm* and *invwarp* FSL utilities, respectively. Quality of spatial transformations was visually assessed by overlay of the contour of standard space participant T1 onto the MNI152 template.

*Tissue-Type Segmentation:* Tissue-probability maps (gray matter (GM), white matter (WM), and cerebrospinal fluid (CSF)) and a tissue-type segmentation were generated with FSL FAST. Additionally, a subcortical segmentation/mask was generated with FIRST (Patenaude et al., 2011). The following steps then utilized *fslmaths* to ‘clean-up’ the segmentation and generate images necessary for subsequent preprocessing:

1. Erroneous CSF voxels were removed from the subcortical mask by removing those voxels that were present in the CSF mask.
2. The ‘cleaned-up’ subcortical mask added into the tissue-type segmentation as GM.
3. A single erosion was performed on GM and CSF masks.
4. An eroded WM mask was generated by eroding WM mask 3 times.
5. An MNI template dilated CSF ventricle mask was transformed into participant space and intersected with the eroded CSF mask to create a participant-specific eroded CSF ventricle mask.

[Masks generated in steps 3-5 were used in fMRI Nuisance Regression]

6. The WM/CSF boundaries of the ventricles (which often get erroneously segmented as GM) were removed from the GM mask via the following:
   a. An interface of WM/CSF was generated as the overlap of the dilated versions of the two masks.
   b. The interface was then masked by the dilated template CSF mask from (5) to isolate the interface around the ventricles.
   c. FSL’s *cluster* tool was then used to take the largest contiguous element of that mask (the area around the ventricles), thus removing any residual small clusters that may not have been caught by the masking in (b).
   d. Finally, the inverse of the WM/CSF ventricle mask was applied to the GM mask to remove any GM misclassified voxels at the WM/CSF boundary.

*Registering Parcellations into Native Space:* Cortical brain parcellations were registered into participant native space via the transformations generated in [*Standard Space Registration*] section. These images were then masked by the participant’s GM mask. Further, to ensure complete coverage of participants’ GM by parcellation an iterative procedure of dilation followed by GM re-masking was carried out 3 times. The cortical GM parcellations were then masked by the inverses of the dilated subcortical mask and the native space-generated cerebellar mask (generated with FSL FIRST) to remove any erroneously assigned voxels that may have resulted from the spatial transformations or iterative dilation/re-masking procedure. The final GM parcellations were then dilated once (to be used in fMRI preprocessing).

**fMRI preprocessing**

*Dicom import:* See [T1 preprocessing: Dicom Import].

*Distortion Correction*: FSL’s *topup* and was used for correction of susceptibility induced distortions, by utilizing 3 pairs of opposite phase-encoding (AP-PA) spin-eco field maps to estimate a susceptibility-induced off-resonance field (Andersson et al., 2003) that was used to correct the fMRI data via *applytopup.*

*Motion Correction:* FSL’s mcFLIRT was used to correct for motion via affine registration to the mean volume (Jenkinson et al., 2002).

*Registration of T1 to fMRI:* A mean volume was created from the motion corrected data and nonbrain tissue was removed via FSL *bet* -R. The following steps describe the stepwise registration of each participant’s T1 to his/her brain-extracted mean fMRI volume:

1. FSL FLIRT linear 6 degrees of freedom registration of T1 to mean fMRI.
2. Apply transformation from (1) to the WM mask.
3. The fMRI mean is then registered to the T1 and WM mask from (1) and (2) with FLIRT boundary-based registration cost function.
4. The transformations from (1) and (3) are then concatenated to produce a single affine transformation from native T1 to native fMRI.

*Registration of Other Images:* The transformation generated in the previous step is then used to register the following onto fMRI native space: brain mask, tissue-type masks, and parcellations.

*Intensity Normalization:* fMRI data were normalized to a global 4D mean of 1000 using *fslmaths -ing* option.

*Nuisance Regression:* For these data a 3-step nuisance regression procedure was utilized. First, the Independent Component Analysis-Automatic Removal of Motion Artifacts (ICA_AROMA) (Pruim et al., 2015) was used to regress out motion related signal while preserving the temporal degrees of freedom in the data, which is necessary for the time-varying analyses employed. Second, the anatomical component-based noise correction method (aCompCor) (Behzadi et al., 2007) was implemented in Matlab and was used to regress out the first 5 principal components of WM and of ventricular CSF signal obtained from the fMRI space-eroded masks. Finally, global signal was regressed from the fMRI data.

*Demean and Detrend:* Each voxel across time was mean-centered and a linear trend was removed with the Matlab *detrend* function.

*Bandpass Filtering:* A first order Butterworth filter (0.009 – 0.08 Hz) was applied to the data via the Matlab butter and filtfilt functions.

*Parcellation timeseries:* The final fMRI voxel time courses were then averaged for each region of interest (ROI) in the parcellation, to obtain the timeseries that were used to generate static and time-varying functional connectivity matrices.
